# DNAmFitAge: Biological Age Indicator Incorporating Physical Fitness

**DOI:** 10.1101/2022.03.21.22272043

**Authors:** Kristen M. McGreevy, Zsolt Radak, Ferenc Torma, Matyas Jokai, Ake T. Lu, Daniel W. Belsky, Alexandra Binder, Luigi Ferrucci, Riccardo E. Marioni, Simon Cox, Michael Kobor, David L. Corcoran, Steve Horvath

## Abstract

Physical fitness is a well-known correlate of health and the aging process. DNA methylation (DNAm) data lend themselves for estimating chronological and biological age through epigenetic clocks. However, current epigenetic clocks did not yet use measures of mobility, strength, lung, or endurance physical fitness parameters in their construction. Here, we develop blood DNAm biomarkers for fitness parameters gait speed (walking speed), hand grip strength, forced expiratory volume in one second (FEV1), and maximal oxygen uptake (VO2max). We then use these DNAm biomarkers to construct DNAmFitAge, a new biological age indicator that incorporates physical fitness with epigenetic mortality risk estimators. Adjusting DNAmFitAge for chronological age generates a novel measure of epigenetic age acceleration, FitAgeAcceleration, which is informative for physical activity level (p=1.2E-12), mortality risk (p=5.9E-13), coronary heart disease risk (p=0.0051), comorbidities (p=9.0E-9), and disease-free status (p=1.1E-6) across several large validation datasets. These newly constructed DNAm biomarkers and DNAmFitAge provide researchers and physicians a new method to incorporate physical fitness into epigenetic clocks and emphasizes the effect of lifestyle on the aging process.

## Introduction

Physical fitness declines with aging and is well known to correlate to health [1]. This decline is evident in reduced function in specific organs, like lungs [2], and in performance tests of strength [3, 4] or aerobic capacity [5, 6]. The rate of this decline varies between individuals [7-10], and those who preserve physical fitness as they age are at lower risk for a range of diseases and tend to live longer lives [11-16]. At the molecular level, changes in fitness and related indices of functional capacity correlate with changes in molecular signs of decline thought to reflect underlying biological processes of aging [17-19]. Measures of fitness may therefore provide a window into biological aging [20]. However, direct measurement of fitness parameters can be challenging, requiring in-person data collection by trained personnel with specialized equipment [21, 22]. Furthermore, fitness measurements are not possible for studies with remote data collection or those conducted with stored biospecimens. To enable such studies to quantify fitness, we developed blood DNAm biomarkers of fitness parameters spanning mobility, strength, lung, and cardiovascular fitness and use these to construct a novel indicator of fitness-based biological age, DNAmFitAge.

Three lines of evidence support a focus on DNAm to develop biomarkers of fitness and aging-related changes in fitness. First, aging is reflected in DNAm changes; thousands of sites across the genome change methylation states as organisms grow older, enabling construction of high-precision algorithms to predict age [23-26]. These are collectively known as epigenetic clocks, and a large body of literature demonstrates these clocks predict lifespan [27, 28], are associated with age-related conditions [28-30], and are reflective of one’s biological age [27, 31]. Second, prediction of aging-related morbidity, disability, and mortality by DNAm biomarkers is enhanced by the incorporation of physiological data into prediction algorithms [27, 28, 32]. This suggests utility in including physical fitness in DNAm biomarkers, however, current DNAm biomarkers do not use fitness parameters in their construction. Third, there is emerging evidence that epigenetic clocks are sensitive to lifestyle factors [33] and that individual differences in fitness parameters are reflected in DNAm data [34-37], but it was unknown if fitness parameters could be estimated using blood DNAm levels.

Here, we develop blood DNAm biomarkers of four fitness parameters: gait speed (walking speed), handgrip strength, forced expiratory volume in 1 second (FEV1; an index of lung function), and maximal oxygen uptake (VO2max; a measure of cardiorespiratory fitness). We then use these biomarkers to develop the novel DNAm fitness-related biological age indicator, DNAmFitAge which quantifies the relationship between physical fitness and biological aging processes. This novel measure incorporates mortality risk with strength, mobility, and cardiovascular fitness using blood DNAm biomarkers. Our newly constructed DNAm biomarkers and DNAmFitAge provide researchers and physicians a new method to incorporate physical fitness into epigenetic clocks and emphasizes the effect lifestyle has on the aging process.

## Methods

### Study Cohorts

We analyzed blood DNAm data from three datasets, Framingham Heart Study Offspring cohort (FHS, n = 1830), Baltimore Longitudinal Study on Aging (BLSA, n=820), and novel data (Budapest, n = 307) to develop DNAm biomarkers of fitness parameters. In short, the Framingham Heart Study is a cardiovascular study which followed adults from Massachusetts starting in 1948 [38]. The Baltimore Longitudinal Study of Aging (BLSA) began in 1958 studying healthy adults and the aging process [39]. Finally, Budapest is a smaller study (n = 307) measuring physical fitness and DNA methylation in middle to older aged adults. Dataset harmonization was performed to join multiple datasets when variables were on different scales following previously developed methods [40]. In brief, datasets were rescaled to have the same mean and standard deviation by recentering and multiplying by the ratio of standard deviations.

We conducted validation analysis in an independent group of six additional datasets: two Lothian Birth Cohorts: LBC1921 (n = 692) and LBC1936 (n = 2797), CALERIE (n = 578), InChianti (n = 924), Jackson Heart Study (JHS, n = 1746), and Women’s Health Initiative (WHI, n = 2117). Descriptive statistics of each dataset are presented in **Table 1**. Full study descriptions for validation datasets have previously been published [41-48].

**Table 1.** Descriptive Statistics for Each Dataset

|  | FHS | BLSA | Budapest | LBC1921 | LBC1936 | CALERIE | InChianti | JHS | WHI |
| --- | --- | --- | --- | --- | --- | --- | --- | --- | --- |
| Total Observations | 1830 | 820 | 307 | 692 | 2797 | 578 | 924 | 1746 | 2117 |
| Age mean (sd) | 66.5 (8.80) | 69.2 (13.6) | 60.3 (11.7) | 82.3 (4.31) | 73.6 (3.7) | 39.4 (7.2) | 67.0 (16.6) | 56.2 (12.3) | 65.4 (7.1) |
| < 40 | 0 (0%) | 24 (2.9%) | 8 (2.6%) | 0 (0%) | 0 (0%) | 265 (45.8%) | 100 (10.8%) | 173 (9.9%) | 0 (0%) |
| 40 - 59 | 414 (22.6%) | 178 (21.7%) | 133 (43.3%) | 0 (0%) | 0 (0%) | 313 (54.2%) | 128 (13.9%) | 856 (49.0%) | 525 (24.8%) |
| 60 - 79 | 1262 (69.0%) | 400 (48.8%) | 151 (49.2%) | 410 (59.2%) | 2719 (97.2%) | 0 (0%) | 502 (54.3%) | 691 (39.6%) | 1589 (75.1%) |
| 80+ | 154 (8.4%) | 218 (26.6%) | 15 (4.9%) | 282 (40.8%) | 78 (2.8%) | 0 (0%) | 194 (21.0%) | 26 (1.5%) | 3 (0.1%) |
| Sex |  |  |  |  |  |  |  |  |  |
| Males | 797 (43.6%) | 417 (50.9%) | 148 (48.2%) | 291 (42.1%) | 1141 (51.5%) | 178 (30.8%) | 426 (46.1%) | 649 (37.2%) | 0 (0%) |
| Females | 1033 (56.4%) | 403 (49.1%) | 159 (51.8%) | 401 (57.9%) | 1356 (48.5%) | 400 (69.2%) | 498 (53.9%) | 1097 (62.8%) | 2117 (100%) |
| Race |  |  |  |  |  |  |  |  |  |
| White | 1830 (100%) | 572 (69.8%) | 297 (96.7%) | 692 (100%) | 2797 (100%) | 442 (76.5%) | 924 (100%) | 0 (0%) | 1007 (47.6%) |
| Black | 0 (0%) | 216 (26.3%) | 0 (0%) | 0 (0%) | 0 (0%) | 71 (12.3%) | 0 (0%) | 1746 (100%) | 677 (32.0%) |
| Asian | 0 (0%) | 24 (2.9%) | 10 (3.3%) | 0 (0%) | 0 (0%) | unknown | 0 (0%) | 0 (0%) | unknown |
| Other | 0 (0%) | 8 (1.0%) | 0 (0%) | 0 (0%) | 0 (0%) | 65 (11.2%) | 0 (0%) | 0 (0%) | 433 (20.5%) |

### DNAm Fitness Parameter Biomarker Development

We developed DNAm biomarkers for four fitness parameters: gait speed, maximum handgrip strength (Gripmax), forced expiratory volume in 1 second (FEV1), and maximal oxygen uptake (VO2max). Gait speed, also known as walking speed, is measured in meters per second [49]. Maximum hand grip strength is a measurement of force taken in kg [8, 12]. FEV1 measures lung function; it is the amount of air forced from the lungs in one second, measured in liters [14]. VO2max is a measure of cardiovascular health and aerobic endurance [6, 50]. It measures the volume of oxygen the body processes during incremental exercise in milliliters used in one minute of exercise per kilogram of body weight (mL/kg/min).

Each fitness DNAm biomarker was developed using LASSO penalized regression with 10-fold cross validation in which the fitness parameters were dependent variables and independent variables were DNAm levels at cytosine-phosphate-guanines (CpG) sites and chronological age. The LASSO-regression method selects a minimum set of covariates that maximizes prediction of the dependent variable from the universe of variables included in the regression. Models were fit separately for men and women in the case of gait speed, gripmax, and FEV1 to allow for sex differences in selected CpG sites. The selected covariates and estimated coefficients were then used to form a prediction algorithm for each fitness parameter. We refer to the DNAm measurements generated by these algorithms as DNAmGaitspeed, DNAmGripmax, DNAmFEV1, and DNAmVO2max. Correlation of each DNAm biomarker with measured fitness values in the training data are displayed in Supplemental Figure 1.

When it came to building the biomarker for VO2max, stratifying by sex was not feasible due to the smaller sample size. This forced us to choose between using sex as a covariate or omitting sex and trusting LASSO to select X chromosome markers that best signify differences between males and females. We chose the latter, and it did. Finally, we present two models for DNAmGaitspeed and DNAmGripmax; one with chronological age and one without chronological age as potential covariates. Removing age as a potential variable for selection in LASSO was performed to remedy high collinearity discovered among these DNAm biomarkers when constructing DNAmFitAge (scatterplot matrix in Supplemental Figure 2).

### DNAm Fitness Parameter Biomarker Validation

We conducted two validation analyses of DNAm biomarkers of fitness parameters using up to five independent datasets. First, we correlated DNAm biomarker values with direct measurements of the fitness parameters. In cases where direct measurement of a fitness parameter was not included in a validation dataset, substitutes were selected. Briefly, gait speed was substituted with a composite leg strength measurement and a composite physical functioning score; FEV1 was substituted by forced expiratory volume (FEV) and VO2max; VO2max was substituted by FEV. Details are reported in Supplemental Note 1.

Second, we evaluated if using our DNAm biomarkers improve estimation of fitness parameters beyond variation explained through age and sex. Direct comparison of our DNAm biomarkers to models only using age and sex as covariates are not possible because they are non-nested models. Instead, we evaluate the significance of the DNAm biomarker as a predictor for each fitness parameter after including age and sex as covariates. Pearson correlations (instead of R squared) are presented in Supplemental Table 1 because they summarize the relationship between estimated and true fitness parameters. The reported p-values indicate the significance of the DNAm biomarker estimate as a predictor for the fitness parameters after including age and sex. The individual-dataset and fixed-effects meta-analysis p-values are calculated across validation datasets with the most relevant variables available in more than one dataset. Specifically, LBC1921 and LBC1936 were used for DNAmGaitSpeed and DNAmFEV1 meta-analysis p-value calculations. LBC1921, LBC1936, CALERIE, and WHI were used for DNAmGripmax. We did not calculate a meta-analysis p-value for DNAmVO2max because only one validation dataset had VO2max measurement.

## DNAmFitAge: Biological Age Estimation

### DNAmFitAge Development

We constructed DNAmFitAge as an indicator of biological age following the methods proposed by Klemera and Doubal [51]. In brief, the Klemera-Doubal model framework stipulates there exists an underlying trait which is unobserved (biological age) which relates to an observable trait (chronological age) and a set of additional variables. This framework posits biological age is centered on chronological age with additional noise. Weighted least squares is used to estimate the relationship of the additional variables with biological age where the weights are formed from correlations of each variable with chronological age.

DNAmFitAge is constructed separately for males and females using four DNAm variables: three of the DNAm fitness biomarkers: DNAmGripmax, DNAmGaitSpeed, and DNAmVO2max, and DNAmGrimAge, a biomarker of mortality risk [28]. We estimate biological age using the TrueTrait function from the WGCNA R package which carries out the Klemera Doubal method described above. Variable weights indicating each variable’s importance for estimating biological age are presented in Table 3A. Pearson’s correlation among original fitness parameters, DNAm biomarkers, and DNAmFitAge in the large training dataset (FHS + BLSA) are displayed in Supplemental Figure 2. Pearson’s correlation of DNAmFitAge to chronological age in training data are presented in panels A and B of Supplemental Figure 3. Models including DNAmFEV1 as a fifth variable were explored, however no improvement in association to physical activity or age-related outcomes were observed; the parsimonious DNAmFitAge model using a subset of the DNAm fitness biomarkers was therefore chosen.

**Table 2.**
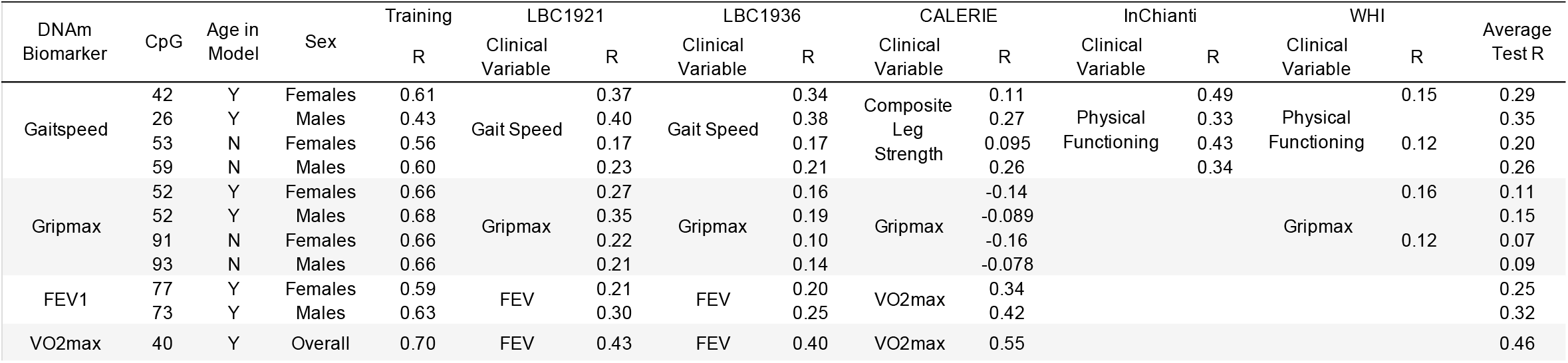
DNAm Fitness Parameter Biomarker Pearson Correlation

**Table 3A.** DNAmFitAge Model Weights

| Variable | Female Weights | Male Weights |
| --- | --- | --- |
| DNAmGripmax | 0.174 | 0.179 |
| DNAmGaitSpeed | 0.228 | 0.159 |
| DNAmVO2max | 0.104 | 0.139 |
| DNAmGrimAge | 0.493 | 0.523 |

**Table 3B.** DNAmFitAge Performance in Validation Datasets

|  |  | Females | Males | Male Model in Females | Female Model in Males |
| --- | --- | --- | --- | --- | --- |
| Training Data | Median Absolute Deviation | 2.7 | 3.0 | 11.9 | 13.5 |
|  | Mean Deviation | 0.0 | 0.0 | -12.2 | 13.1 |
|  | R | 0.923 | 0.925 | 0.925 | 0.922 |
| LBC1921 | Median Absolute Deviation | 3.7 | 4.8 | 11.0 | 14.5 |
|  | Mean Deviation | 0.8 | 1.1 | -11.1 | 13.8 |
|  | R | 0.409 | 0.386 | 0.404 | 0.391 |
| LBC1936 | Median Absolute Deviation | 3.2 | 3.4 | 11.6 | 13.3 |
|  | Mean Deviation | 0.0 | 0.2 | -11.9 | 12.9 |
|  | R | 0.635 | 0.635 | 0.647 | 0.624 |
| CALERIE | Median Absolute Deviation | 4.9 | 2.3 | 17.1 | 11.0 |
|  | Mean Deviation | -5.0 | -2.0 | -17.1 | 11.0 |
|  | R | 0.926 | 0.915 | 0.928 | 0.912 |
| InChianti | Median Absolute Deviation | 3.9 | 3.9 | 16.0 | 9.6 |
|  | Mean Deviation | -3.8 | -4.3 | -16.1 | 9.1 |
|  | R | 0.969 | 0.964 | 0.969 | 0.963 |
| JHS | Median Absolute Deviation | 2.9 | 3.4 | 13.6 | 9.2 |
|  | Mean Deviation | -1.6 | -2.8 | -13.9 | 8.6 |
|  | R | 0.937 | 0.917 | 0.940 | 0.914 |
| WHI | Median Absolute Deviation | 3.8 |  | 16.8 |  |
|  | Mean Deviation | -3.4 |  | -16.8 |  |
|  | R | 0.808 |  | 0.812 |  |

Finally, we created FitAgeAcceleration, the age-adjusted estimate of DNAmFitAge formed from taking the residuals after regressing DNAmFitAge onto chronological age. As such, FitAgeAcceleration is uncorrelated with chronological age. FitAgeAcceleration provides an estimate of epigenetic age acceleration, ie how much older or younger a person’s estimated biological age is from expected chronological age. A positive FitAgeAcceleration means biological age is estimated to be older than chronological age. A negative FitAgeAcceleration means biological age is estimated to be younger than chronological age, which is the preferred outcome for a person.

### DNAmFitAge Validation

DNAmFitAge validation analysis consisted of three components: correlating DNAmFitAge to chronological age, testing FitAge Acceleration association with physical activity, and testing FitAge Acceleration association to aging-related variables in the validation datasets. First, the modeling framework posits biological age is centered on chronological age, therefore validation datasets should demonstrate good correlation and general centeredness between DNAmFitAge and chronological age. Both properties would indicate DNAmFitAge can quantify age. Second, DNAmFitAge incorporates fitness, therefore FitAgeAcceleration (age adjusted DNAmFitAge) should relate to physical activity and physical functioning. These relationships would indicate DNAmFitAge relates to fitness. Third, DNAmFitAge provides insight to the aging process through a fitness paradigm, therefore FitAgeAcceleration should relate to aging-related phenotypes.

We correlate DNAmFitAge with chronological age for males and females because (1) we cannot directly measure biological age, (2) chronological age is not used when forming DNAmFitAge estimates, and (3) the modeling framework posits biological age is centered on chronological age. In addition, because DNAmFitAge is built in males and females separately, we demonstrate what happens when the model is applied to the opposite sex (ie male model in females or female model in males). Median absolute deviation, mean deviation, and Pearson correlation are presented in Table 3B and displayed in Figure 2.

**Figure 1.**
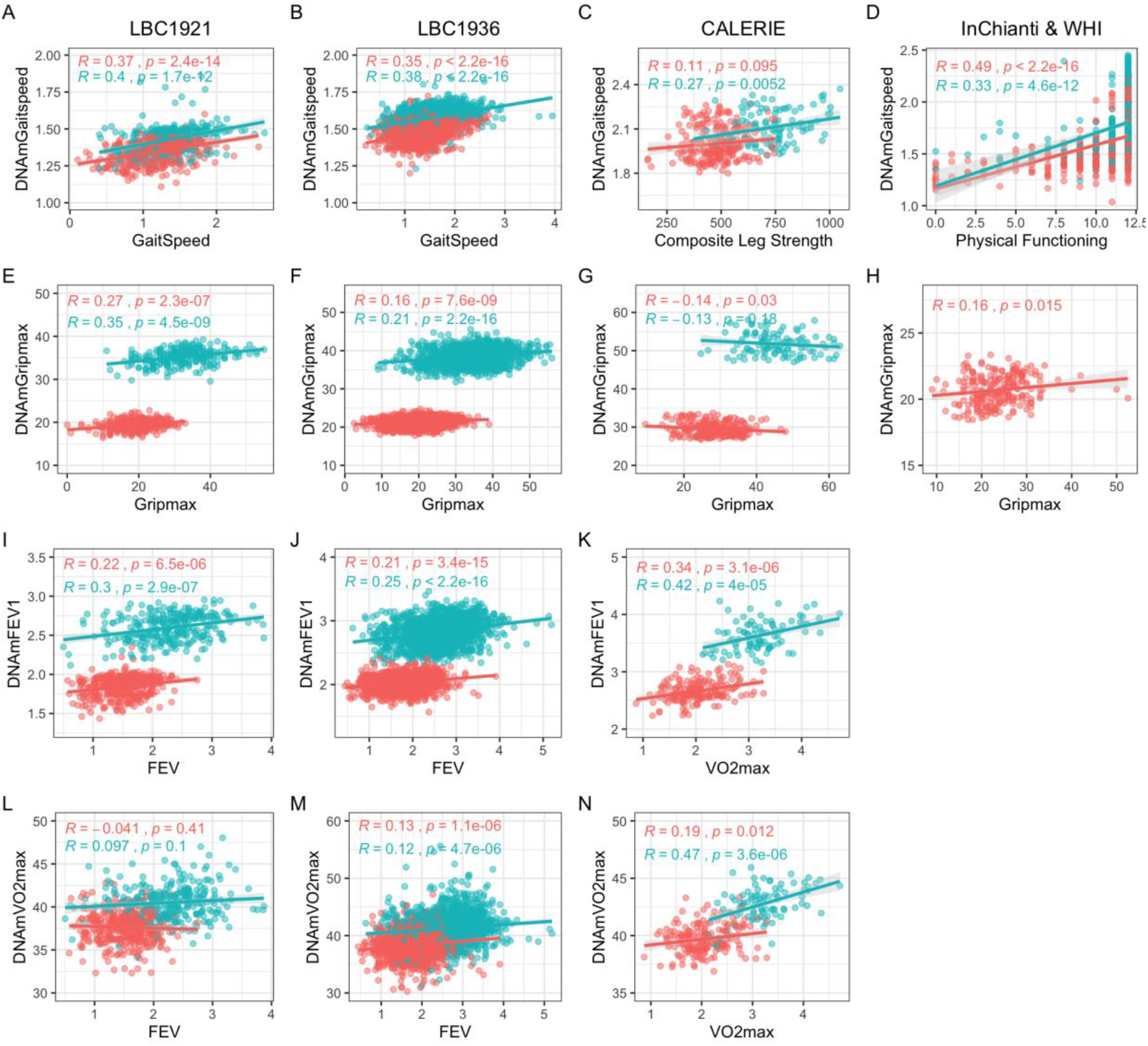
Scatterplots of DNAm fitness biomarker models versus true values in test datasets. Pink indicates females, and blue indicates males. When original variables were unavailable, best alternative variables are plotted against the DNAm fitness estimates. Each panel corresponds to the performance of one DNAm-based model built with chronological age across test datasets displayed with Pearson correlation and p-values. (**A-D)** DNAmGaitspeed, (**E-H)** DNAmGripmax, (**I-K)** DNAmFEV1, (**L-N)** DNAmVO2max. (**A-K)** (DNAmGaitspeed, DNAmGripmax, and DNAmFEV1) were built in each sex separately while (**L-N)** (DNAmVO2max) was built in both sexes jointly. (**D)** displays performance in InChianti dataset, and (**H)** displays performance in WHI dataset.

**Figure 2.**
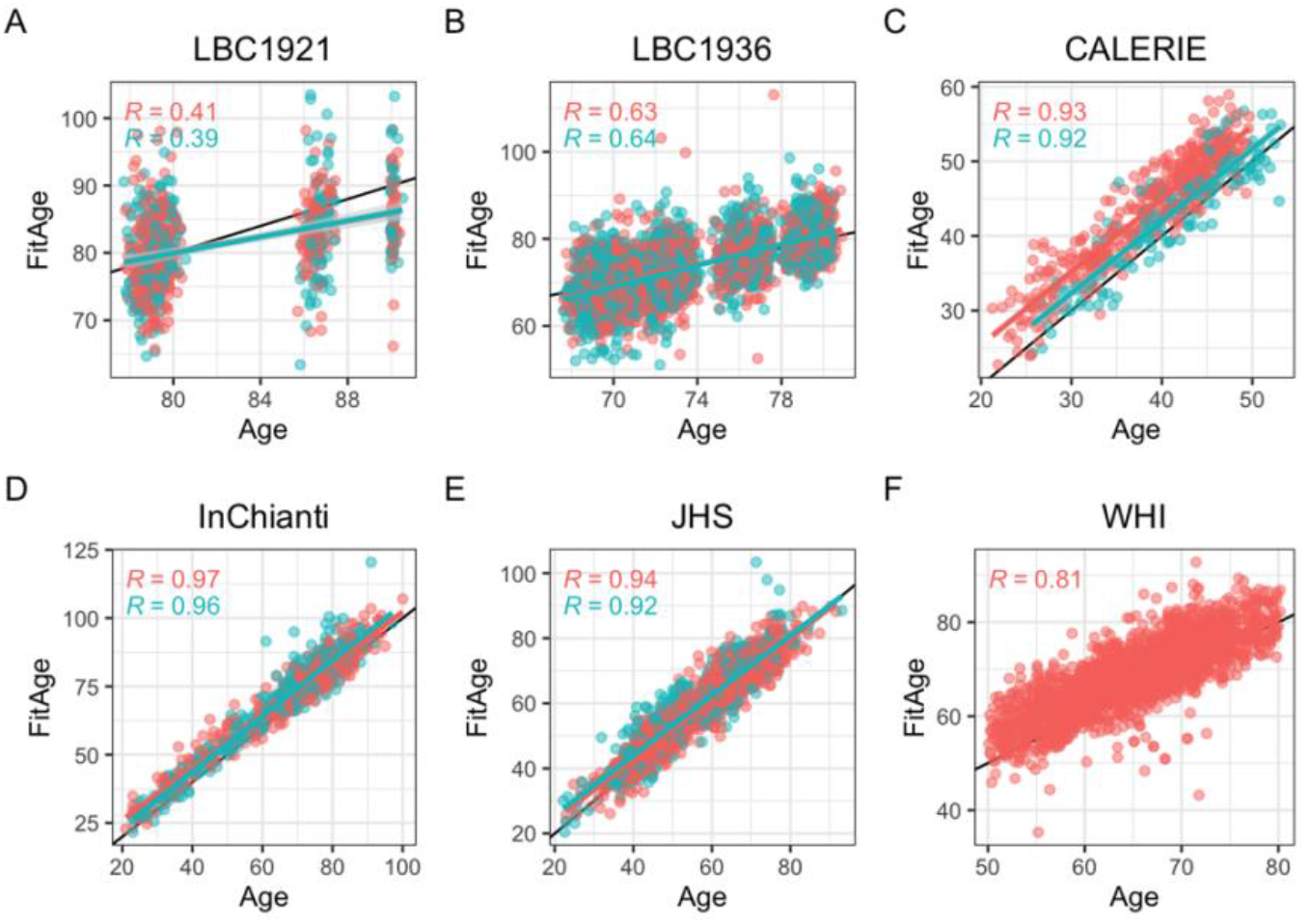
Scatterplots of DNAmFitAge versus age separated by sex. Pink indicates females, and blue indicates males. Each row corresponds to the performance of DNAmFitAge across datasets displayed with Pearson correlation to chronological age and corresponding p-values. **(A-G)** DNAmFitAge models applied to the same sex it was built in (ie DNAmFitAge built for females tested in females and DNAmFitAge built for males tested in males). DNAmFitAge is centered on chronological age with high correlation in test sets. (**H-N)** DNAmFitAge models applied to the opposite sex it was built in (ie DNAmFitAge built for females tested in males and DNAmFitAge built for males tested in females). Females are estimated to be older than they are, and males are estimated to be younger than they are.

We tested for associations between physical activity or physical functioning in low to intermediate physically fit individuals with FitAgeAcceleration, DNAm fitness parameter biomarkers, and other DNAm biomarkers known to relate to physical health. We restricted our analysis to people of low to intermediate fitness to determine if FitAgeAcceleration is more sensitive to small improvements in fitness compared to other current DNAm biomarkers. In addition, this separation captures low to average physically active individuals in each dataset. Furthermore, research suggests any exercise compared to none is beneficial to health [52], and we hope DNAmFitAge may serve as a tool to motivate starting an exercise regimen at any level. In short, LBC1921, LBC1936, and JHS measure physical activity, and WHI and InChianti measure physical functioning. Higher values of any variable indicate more activity or better physical functioning. Other DNAm biomarkers which relate to physical health include DNAmGrimAge [28], DNAmPhenoAge [27], DNAmPAI-1 [28], and DNAmGDF-15 [28]. See Supplemental Note 1 for a thorough description of physical activity variables and inclusion criteria.

We tested DNAmFitAge associations to multiple aging-related variables in validation datasets. Specifically, we conducted regression analysis of physical activity, time-to-death, time-to-coronary-heart-disease (CHD), the count of age-related conditions (arthritis, cataract, cancer, CHD, CHF, emphysema, glaucoma, lipid condition, osteoporosis, and type 2 diabetes), age at menopause, and cancer hypertension, type-2 diabetes, and disease-free status.

Time-to-event outcomes were analyzed using Cox regression to estimate hazard ratios (HR); continuous outcomes were analyzed using linear regression; dichotomous outcomes were analyzed using logistic regression to estimate odds ratios (OR); and ordinal outcomes were analyzed using multinomial regression to estimate OR. Linear regression models with person-level random intercepts were implemented in R using the lmer function. Logistic regression models were estimated using generalized estimating equations with the R function gee. Multinomial models were implemented using R function multinom.

We combine results across validation studies using fixed effect models or Stouffer’s meta analysis method using the metafor R function. Fixed effect models use the inverse variance to weight estimates, and Stouffer’s method uses the square root of the sample size to weight estimates. The latter is used when harmonization across cohorts was challenging; such as with physical activity variables, the number of age-related conditions, disease free status, and age at menopause. Forest plots evaluating FitAgeAcceleration hazard ratios or coefficients in models adjusted for age and sex are displayed in Figure 4 and Supplemental Table 2.

**Figure 3.**
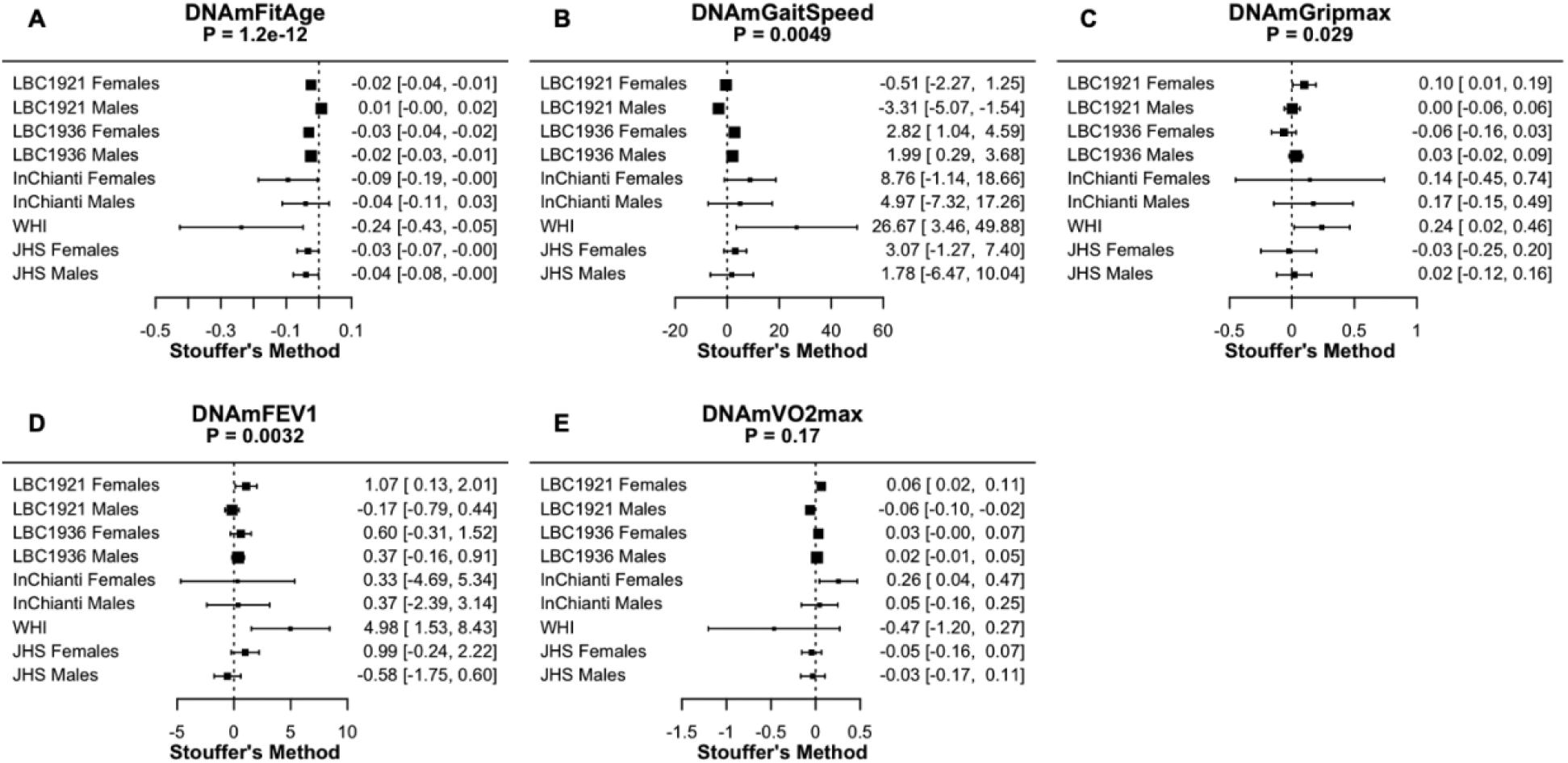
Meta-analysis forest plots for DNAmFitAge and DNAm fitness parameters relationship to physical activity or physical functioning in people with low to intermediate physical activity. Each panel reports the Stouffer’s meta-analysis p-value for combining coefficients across dataset cohorts after adjusting for chronological age. **(A)** DNAmFitAge, **(B)** DNAmGaitspeed, **(C)** DNAmGripmax, **(D)** DNAmFEV1, and **(E)** DNAmVO2max. DNAmFitAge, DNAmGaitSpeed, DNAmGripmax, and DNAmFEV1 are predictive of physical activity in low to intermediately active individuals.

**Figure 4.**
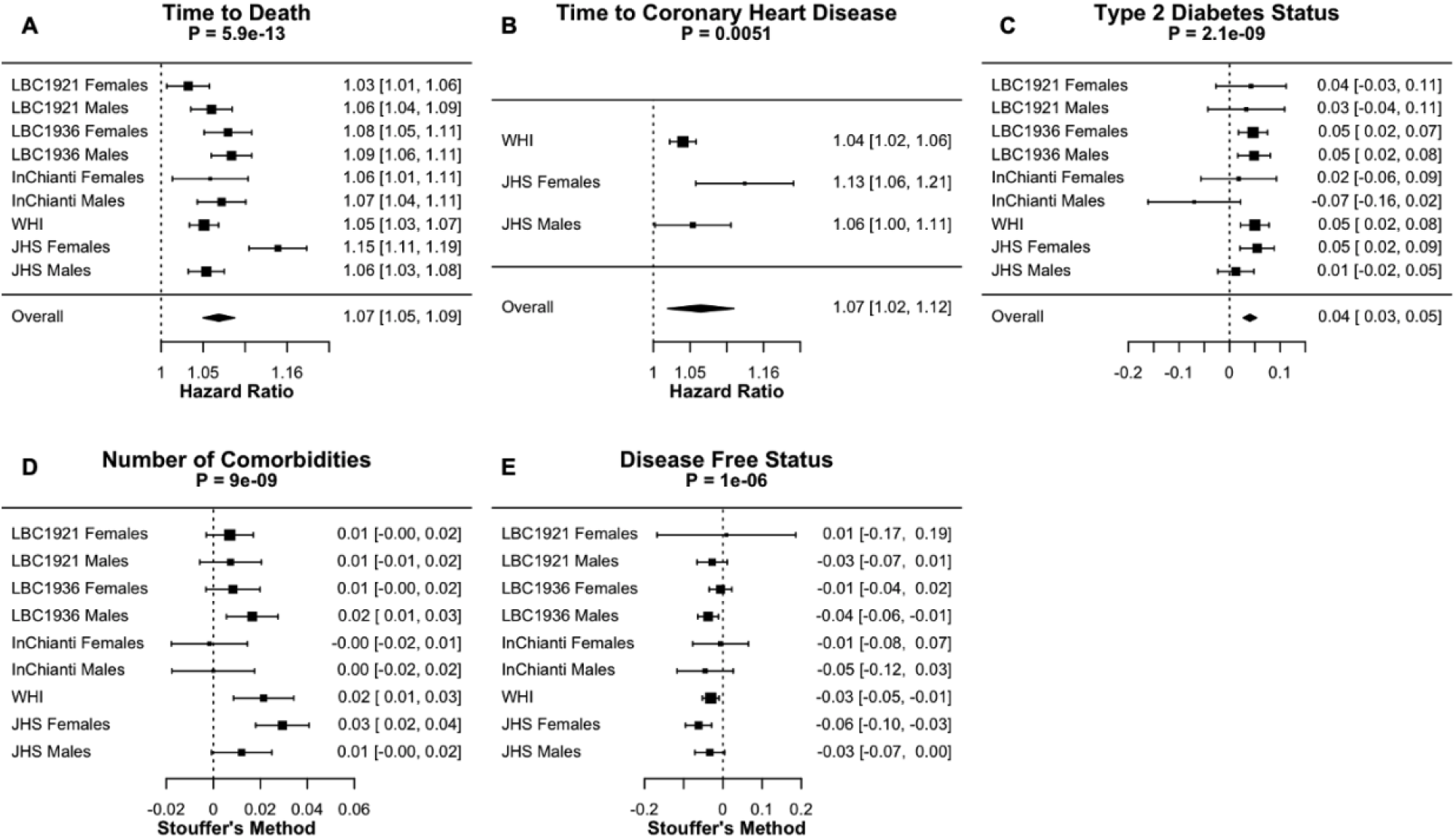
Meta-analysis forest plots for FitAgeAcceleration to age-related conditions adjusted for age and sex. Each panel reports a meta analysis forest plot for combining hazard ratios or regression coefficients across dataset cohorts. (**A**) Time-to-death, (**B**) time-to-coronary heart disease, (**C**) type 2 diabetes, (**D**) comorbidity count, and (**E**) disease free status. Meta-analysis p-values are displayed in the header of each panel. Fixed effects models were used for (**A-C**) and Stouffer’s method was used for (**D, E**).

### GREAT Analysis

We applied the GREAT analysis software tool to provide biological insight to the CpG loci used in constructing our DNAm biomarker estimates. GREAT analyzes genes within and nearby the genomic region covered by the CpGs; it performs a binomial test (over genomic regions) using a whole genome background. We performed the enrichment based on default settings (Proximal: 5.0 kb upstream, 1.0 kb downstream, plus Distal: up to 1,000 kb). We report nominal, Bonferroni, and FDR p-values for gene, biological, cellular, and molecular function in Table 6.

## Results

### DNAm Fitness Parameter Biomarker Models

The number of CpG loci selected through LASSO for each DNAm model are presented in Table 2. Between 40 and 77 CpG loci were needed in addition to chronological age to estimate each fitness parameter. Without age as a covariate in the DNAm biomarker estimates, more CpG loci were needed to achieve similar precision; between 53 and 93. DNAmVO2max model includes several CpG loci on the X chromosome, likely capturing sex effects.

The DNAm fitness parameter biomarkers had modest correlation with direct fitness parameters. Average correlations ranged from 0.07-0.46 (scatterplots are shown in **Figure 1**; Pearson correlations are reported in **Table 2**). DNAmGripmax in males and females had moderate correlations in validation datasets but do not perform well in CALERIE. We hypothesize this may be due in part from the stringent enrollment criteria: free of chronic disease, non-obese, and relatively young. Correlation of DNAmVO2max to FEV in LBC1921 and LBC1936 was weak within each sex; however varying correlations have been described in literature [53-56]. Reported correlations between VO2max and FEV vary from 0 to 0.5, likely because VO2max is a measure of cardiovascular health whereas FEV is a measure of lung volume. Correlation of DNAmVO2max to VO2max in CALERIE, the one validation dataset with the direct fitness parameter, has good correlation overall and within sex (overall R=0.55, female R=0.19, male R=0.47).

The DNAm biomarkers improve estimation of fitness parameters beyond what is explained through age and sex in many validation datasets (Supplemental Table 1). Interestingly, DNAmGaitspeed and DNAmGripmax built without chronological age have lower correlation with true fitness parameters compared to the models built with chronological age. However, the DNAm biomarkers built without chronological age explain more additional variation in fitness parameters compared to the age-included DNAm biomarkers. This suggests the DNAm biomarkers capture different information than age and sex for understanding fitness parameters.

DNAmGaitspeed, DNAmFEV1, and DNAmVO2max are predictive of mortality, and DNAmGaitspeed and DNAmFEV1 are strong predictors for number of comorbidities in the validation datasets. Relationship of each DNAm fitness biomarker with time-to-death, type 2 diabetes, number of comorbidities, and disease free status after adjusting for age are displayed in Supplemental Figure 4. Relationship of DNAm biomarkers to physical activity are explored alongside DNAmFitAge below.

### DNAmFitAge

DNAmFitAge was built in each sex using DNAmGripmax, DNAmVO2max, DNAmGaitSpeed, and DNAmGrimAge. The models had similar weights in males and females and had strong correlation with and is generally centered on chronological age. DNAmGripmax, DNAmVO2max, and DNAmGaitspeed contributed around 50% to estimating DNAmFitAge in each sex, and DNAmGrimAge contributed the remaining 50% (Table 3A). Weights were very similar in males (13.9 - 17.9%), and there was slightly more variation in females (10.4 - 22.4%). This may suggest that DNAmGaitSpeed was more influential in estimating biological age in females than males. DNAmFitAge had strong correlation to chronological age in validation datasets, and the lower correlation in LBC1921 (R = 0.38) and LBC1936 (R = 0.68) can be attributed to the small age range they cover. LBC1921 ages ranged from 77 to 90 and LBC1936 ages ranged from 67 to 80. The average Pearson R across all validation datasets was 0.77, and the average R excluding LBC cohorts was 0.92. In addition, each validation dataset had low median absolute deviation (average difference from chronological age to biological age) ranging from 2.3 to 4.9 years. Overall, DNAmFitAge had high correlation with chronological age and great reproducibility across the validation datasets. In addition, the validation datasets span a large range of chronological age-from 21 (CALERIE) to 100 (InChianti). Reproducibility across a wide span of ages demonstrate DNAmFitAge’s calibration across a wide adult age range.

Applying each DNAmFitAge model to the opposite sex shows strong correlation with age but with substantial over and underestimation of age in females and males, respectively. Females were estimated to be older than they truly are using the DNAmFitAge model built in males (mean deviation -12.2 years) (Table 3B, Supplemental Figure 3). Males were estimated to be younger than they truly are using the DNAmFitAge model built in females (mean deviation +13.1 years). Over and under estimation can be explained by universal differences in fitness parameters by sex. Females tended to have lower fitness parameters compared to males. Hence males were predicted to be younger than they are using the female DNAmFitAge model because larger values of DNAmGaitSpeed, DNAmGripmax, or DNAmVO2max would indicate stronger (or more physically fit) females. To provide an idea of DNAm fitness biomarker values which correspond to fit/young DNAmFitAge compared to unfit/ old DNAmFitAge, we provide reference values within age and sex categories in Table 4. Overall, higher or more physically fit values of DNAmGaitspeed, DNAmGripmax, or DNAmGaitspeed corresponded to younger estimated biological ages in males and females.

**Table 4.** Reference DNAm Fitness Parameter Values for Fit (FitAge Acceleration <=-5 yrs) and Unit (FitAge Acceleration >=+5 yrs) Individuals

| Age | Females |  |  |  |  |  |  |  |
| --- | --- | --- | --- | --- | --- | --- | --- | --- |
|  | DNAMGaitspeed |  | DNAMGripmax |  | DNAMVO2max |  | DNAMGrimAge |  |
|  | Fit | Unfit | Fit | Unfit | Fit | Unfit | Fit | Unfit |
| < 40 | 2.1 | 2.0 | 34.6 | 30.5 | 42.8 | 40.1 | 37.1 | 40.9 |
| 40-59 | 1.9 | 1.7 | 31.3 | 26.9 | 39.2 | 37.9 | 49.2 | 60.5 |
| 60-79 | 1.7 | 1.5 | 28.8 | 22.4 | 37.6 | 36.1 | 63.2 | 72.4 |
| 80+ | 1.6 | 1.3 | 23.9 | 19.1 | 37.0 | 35.4 | 74.7 | 81.8 |
| Age | Males |  |  |  |  |  |  |  |
|  | DNAMGaitspeed |  | DNAMGripmax |  | DNAMVO2max |  | DNAMGrimAge |  |
|  | Fit | Unfit | Fit | Unfit | Fit | Unfit | Fit | Unfit |
| < 40 | 2.1 | 1.8 | 49.3 | 43.8 | 45.1 | 44.9 | 34.8 | 52.9 |
| 40-59 | 1.9 | 1.7 | 46.6 | 42.5 | 43.9 | 42.3 | 47.5 | 60.1 |
| 60-79 | 1.7 | 1.5 | 41.3 | 36.8 | 43.1 | 39.5 | 68.0 | 77.9 |
| 80+ | 1.6 | 1.3 | 39.3 | 32.0 | 41.3 | 37.7 | 78.0 | 86.7 |

### DNAmFitAge Relationship to Physical Activity

FitAgeAcceleration, DNAmGaitspeed, DNAmGripmax, and DNAmFEV1 have associations in the expected direction with physical activity in low to intermediate physically active individuals. Coefficients indicate the effect on physical activity for a one unit increase in each DNAm surrogate marker after adjusting for chronological age within each sex (Table 5, Figure 3). The relationship to DNAmFitAge is as expected; someone with a higher FitAgeAcceleration has an estimated biological age that is older than expected, which corresponds to lower physical activity or physical functioning (Table 4). Similarly, men and women with a faster DNAmGaitspeed, stronger DNAmGripmax, and larger DNAmFEV1 are more physically active when holding age constant. In conclusion, men and women who were more active showed correspondingly ‘fitter’ values of FitAgeAcceleration and the DNAm fitness biomarkers. Additionally, DNAmFitAge (Stouffer p-value = 1.2 E -12) marginally outperforms current DNAm biomarkers when comparing meta-analysis p-values; improvement of DNAmFitAge compared to DNAmGrimAge (p-value = 1.0 E-11) is marginal, however the improvement compared to DNAmPhenoAge (p-value = 1.9 E-5), DNAmGDF-15 (p-value = 1.1 E-8), and DNAmPAI-1 (p-value = 1.6 E -9) is more pronounced. In addition, DNAmFitAge, which provides an indicator of biological age, may provide a more interpretable aging biomarker compared to DNAmGrimAge, which provides a measurement of lifespan. These comparisons demonstrate DNAmFitAge can capture the relationship to physical activity and can provide an improvement to the arsenal of current DNAm biomarkers.

**Table 5.**
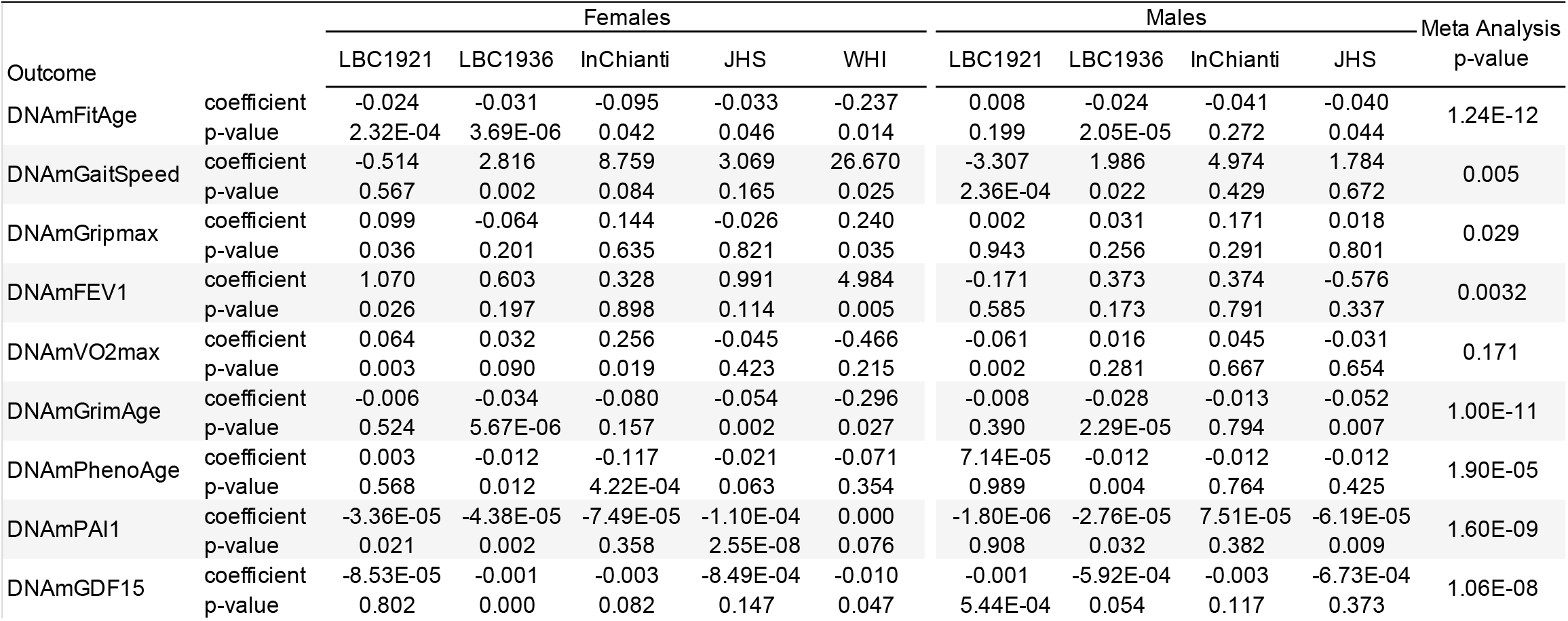
Association of DNAm Biomarkers to Physical Activity and Physical Functioning in People with Low to Intermediate Activity Levels

| Outcome |  | Females |  |  |  |  | Males |  |  |  | Meta Analysis p-value |
| --- | --- | --- | --- | --- | --- | --- | --- | --- | --- | --- | --- |
|  |  | LBC1921 | LBC1936 | InChianti | JHS | WHI | LBC1921 | LBC1936 | InChianti | JHS |  |
| DNAmFitAge | coefficient | -0.024 | -0.031 | -0.095 | -0.033 | -0.237 | 0.008 | -0.024 | -0.041 | -0.040 | 1.24E-12 |
|  | p-value | 2.32E-04 | 3.69E-06 | 0.042 | 0.046 | 0.014 | 0.199 | 2.05E-05 | 0.272 | 0.044 |  |
| DNAmGaitSpeed | coefficient | -0.514 | 2.816 | 8.759 | 3.069 | 26.670 | -3.307 | 1.986 | 4.974 | 1.784 | 0.005 |
|  | p-value | 0.567 | 0.002 | 0.084 | 0.165 | 0.025 | 2.36E-04 | 0.022 | 0.429 | 0.672 |  |
| DNAmGripmax | coefficient | 0.099 | -0.064 | 0.144 | -0.026 | 0.240 | 0.002 | 0.031 | 0.171 | 0.018 | 0.029 |
|  | p-value | 0.036 | 0.201 | 0.635 | 0.821 | 0.035 | 0.943 | 0.256 | 0.291 | 0.801 |  |
| DNAmFEV1 | coefficient | 1.070 | 0.603 | 0.328 | 0.991 | 4.984 | -0.171 | 0.373 | 0.374 | -0.576 | 0.0032 |
|  | p-value | 0.026 | 0.197 | 0.898 | 0.114 | 0.005 | 0.585 | 0.173 | 0.791 | 0.337 |  |
| DNAmVO2max | coefficient | 0.064 | 0.032 | 0.256 | -0.045 | -0.466 | -0.061 | 0.016 | 0.045 | -0.031 | 0.171 |
|  | p-value | 0.003 | 0.090 | 0.019 | 0.423 | 0.215 | 0.002 | 0.281 | 0.667 | 0.654 |  |
| DNAmGrimAge | coefficient | -0.006 | -0.034 | -0.080 | -0.054 | -0.296 | -0.008 | -0.028 | -0.013 | -0.052 | 1.00E-11 |
|  | p-value | 0.524 | 5.67E-06 | 0.157 | 0.002 | 0.027 | 0.390 | 2.29E-05 | 0.794 | 0.007 |  |
| DNAmPhenoAge | coefficient | 0.003 | -0.012 | -0.117 | -0.021 | -0.071 | 7.14E-05 | -0.012 | -0.012 | -0.012 | 1.90E-05 |
|  | p-value | 0.568 | 0.012 | 4.22E-04 | 0.063 | 0.354 | 0.989 | 0.004 | 0.764 | 0.425 |  |
| DNAmPAI1 | coefficient | -3.36E-05 | -4.38E-05 | -7.49E-05 | -1.10E-04 | 0.000 | -1.80E-06 | -2.76E-05 | 7.51E-05 | -6.19E-05 | 1.60E-09 |
|  | p-value | 0.021 | 0.002 | 0.358 | 2.55E-08 | 0.076 | 0.908 | 0.032 | 0.382 | 0.009 |  |
| DNAmGDF15 | coefficient | -8.53E-05 | -0.001 | -0.003 | -8.49E-04 | -0.010 | -0.001 | -5.92E-04 | -0.003 | -6.73E-04 | 1.06E-08 |
|  | p-value | 0.802 | 0.000 | 0.082 | 0.147 | 0.047 | 5.44E-04 | 0.054 | 0.117 | 0.373 |  |

**Table 6.** GREAT CpG Annotation

| Genes | Observed Regions | Fold Enrichment | Nominal p-value | Bonferroni p-value | FDR Q-value |
| --- | --- | --- | --- | --- | --- |
| HLA-G | 5 | 44.4 | 1.36E-07 | 0.0025 | 0.0025 |
| CD3EAP | 3 | 279.8 | 2.03E-07 | 0.0038 | 0.0019 |
| ERCC2 | 3 | 148.7 | 1.34E-06 | 0.025 | 0.0083 |
| ZNRD1 | 4 | 50.3 | 1.55E-06 | 0.029 | 0.0072 |
| **Biological** |  |  |  |  |  |
| regulation of ketone metabolic processes | 6 | 21 | 5.77E-07 | 0.0074 | 0.0074 |
| antigen processing and presentation via MHC class I, TAP independent | 6 | 17.7 | 1.55E-06 | 0.02 | 0.01 |
| negative regulation of dendritic cell differentiation | 5 | 22.4 | 3.77E-06 | 0.049 | 0.016 |
| **Cellular** |  |  |  |  |  |
| MHC protein complex | 12 | 21.6 | 1.02E-12 | 1.69E-09 | 1.69E-09 |
| integral component of lumenal side of endoplasmic reticulum membrane | 10 | 11.7 | 2.45E-08 | 4.07E-05 | 2.04E-05 |
| intrinsic component of endoplasmic reticulum membrane | 29 | 3.3 | 6.25E-08 | 0.0001 | 3.47E-05 |
| integral component of endoplasmic reticulum membrane | 28 | 3.2 | 1.35E-07 | 0.00023 | 5.63E-05 |
| MHC class I protein complex | 6 | 22.4 | 4.02E-07 | 0.00067 | 0.00013 |
| MHC class II protein complex | 6 | 20.8 | 6.09E-07 | 0.001 | 0.00017 |
| DNA-directed RNA polymerase I complex | 7 | 11 | 4.62E-06 | 0.0077 | 0.0011 |
| MMXD complex | 4 | 35.3 | 6.23E-06 | 0.01 | 0.0013 |
| **Molecular Function** |  |  |  |  |  |
| peptide antigen binding | 9 | 12.9 | 5.56E-08 | 0.00023 | 0.00023 |
| MHC class II receptor activity | 5 | 23 | 3.32E-06 | 0.014 | 0.0069 |
| DNA-directed 5'-3' RNA polymerase activity | 11 | 5.9 | 3.98E-06 | 0.017 | 0.0055 |
| antigen binding | 15 | 4.2 | 4.87E-06 | 0.02 | 0.0051 |
| 5'-3' RNA polymerase activity | 11 | 5.7 | 5.39E-06 | 0.022 | 0.0045 |

### FitAgeAcceleration in Age-related Conditions

FitAgeAcceleration as an epigenetic age acceleration measure was an important predictor of mortality, coronary heart disease, and other age-related conditions. Cox Proportional Hazard models demonstrated FitAgeAcceleration is a strong predictor for time-to-death (p = 5.9 E-13) and time-to-coronary heart disease (p = 0.0051). FitAgeAcceleration had an overall hazard ratio of 1.07 (1.05, 1.09) (Figure 4). Thus, a FitAgeAcceleration value of 10 years was associated with almost doubling the mortality risk compared to the average person of the same age and sex (1.07^10 = 1.97 risk). Similarly, increase in FitAgeAcceleration corresponds to more comorbidities (p = 9.0E-9), hypertension (p = 7.3E-5), and earlier age at menopause (p = 6.6 E-9) (Figure 4, Supplemental Table 2). A lower FitAgeAcceleration was associated with disease free status (p= 1.1 E-6) and lower cholesterol (p = 0.0005) (Supplemental Table 2).

Each of these associations were in the expected direction, as someone who had a low FitAgeAcceleration had a biological age estimate that was younger than their chronological age. Hence, people who were estimated to be more ‘physically fit’ had better age-related outcomes. These relationships demonstrate epigenetic age acceleration can be well explained through DNAm fitness parameter biomarkers, and that FitAgeAcceleration provides a practical tool for relating fitness to the aging process.

FitAgeAcceleration was also explored for explaining information beyond what is captured through DNAmGrimAge and the age-adjusted measure AgeAccelGrim with age-related conditions. DNAmFitAge is built using GrimAge, and GrimAge is known to be associated with age-related conditions. Therefore, FitAgeAcceleration is compared in two sets of models; one adjusts for age and sex, and the other adjusts for age, sex, and AgeAccelGrim. FitAge Acceleration improves mortality risk estimation after including AgeAccelGrim in the LBC1936 and InChianti datasets when comparing LRT p-values (Supplemental Table 3). Overall, our results indicate FitAgeAcceleration is informative for mortality risk and may act as a supplement (not replacement) to AgeAccelGrim.

### Functional CpG Annotation

The 971 CpG genomic locations used to construct the DNAm biomarker estimates were analyzed using the GREAT software tool. Twelve gene sets were found below the FDR of 0.05, and four surpassed the more stringent Bonferroni correction: histocompatibility antigen (HLA-G), CD3e epsilon associated protein (CD3EAP), excision repair 2 (ERCC2), and zinc ribbon domain containing 1 (ZNRD1) (Table 6). CD3EAP is located between ERCC1 and ERCC2 on chromosome 19 (22K base pairs apart), an area involved in DNA repair and apoptosis.

Interestingly, CD3EAP expression was observed to change in skeletal muscle following 12 weeks of endurance exercise [57]. However, additional research examining CD3EAP’s relationship to physical fitness was not found in our literature search.

The top biological processes included regulation of cellular ketone metabolism, antigen processing via MHC class I, and regulation of dendritic cell differentiation. Ketone metabolism is required for energy during exercise, and peripheral blood dendritic cells have been shown to increase following intense physical activity [58]. Both biological findings are intriguing and may provide direction for studying modifiable methylation from fitness parameters. Molecular and cellular processes were found to both relate to MHC protein complexes; this relationship to inflammation-based genes and processes like HLA and MHC may support hypotheses relating physical fitness and systemic inflammation [59]. However, this inflammation relationship may be confounded by the DNA methylation being taken in blood.

## Discussion

DNAm biomarkers have been constructed for blood cell count, age, smoking, and more, however, there were not yet DNAm biomarkers for fitness parameters. Our work introduces new DNAm biomarkers for the fitness parameters of grip strength, gait speed, FEV1, and VO2max. These DNAm biomarkers represent new tools for researchers with access to blood samples and interest in epigenetic components to fitness. For example, VO2max measurement requires specialized equipment, trained personnel, and can be unsafe to measure in older adults [21, 22]. However, while DNAmVO2max can be easily measured with a blood draw, it exhibits only moderate correlation with the measured VO2max (on average *r*=0.46 in the validation data). Our DNAm biomarker fitness parameter biomarkers are not intended to replace true physical fitness measurements. Instead, these DNAm biomarker estimates provide an epigenetic component to evaluating a person’s physical fitness.

DNAm biomarkers have been improved by incorporating phenotypic information, however, DNAm biomarkers had not yet incorporated physical fitness. DNAmFitAge provides researchers a novel indicator of biological age which combines physical fitness and epigenetic health. This biomarker integrates the established DNAm prediction of mortality risk (DNAmGrimAge) with the newly developed DNAm predictions of fitness. Higher values of DNAm fitness biomarkers, which reflect greater physical fitness, correspond to younger estimated biological ages in men and women. Furthermore, DNAmFitAge is associated with physical activity in people of low to intermediate activity levels across five large-scale validation datasets and can outperform some DNAm biomarkers. FitAgeAcceleration is strongly associated with a host of age-related conditions and predicts time-to-death and time-to-CHD across validation datasets. In summary, DNAmFitAge provides an easily interpretable tool to relate physical fitness to biological age, and the age-adjusted version, FitAgeAcceleration, provides a novel measure of epigenetic age acceleration explained through physical fitness.

We acknowledge the following limitations. The DNAm fitness parameter biomarkers have marginal improvement to estimate fitness parameters after including age and sex as covariates in validation datasets. Therefore, the DNAm biomarkers should not *replace* true fitness parameters. Instead, our DNAm fitness biomarkers can supplement direct measurements to understand physical fitness and physiological health from an epigenetic perspective. In addition, our DNAmVO2max biomarker was only validated in one dataset with VO2max; more research is needed to evaluate how our DNAmVO2max biomarker performs across a range of independent datasets. Finally, we studied generally healthy adults and did not explore relationships in samples of athletes or teenagers. Additional research is needed to understand performance in younger, more athletic populations.

Overall, DNAmGaitspeed, DNAmGripmax, DNAmFEV1, DNAmVO2max, DNAmFitAge, and FitAgeAcceleration provide epigenetic components to evaluating a person’s physical fitness and biological age. This research demonstrates biological age can be estimated using DNAm fitness parameter biomarkers which are dependent on exercise lifestyle. DNAmFitAge and FitAgeAcceleration are practical and intuitive tools; physically fit people have a younger DNAmFitAge and younger FitAgeAcceleration, and younger values are associated with more physical activity and better age-related outcomes. Our research suggests exercise and stronger fitness parameters are protective to DNAmFitAge in both sexes, and additional research should explore if DNAmFitAge and FitAgeAcceleration are sensitive to exercise interventions.

## Data Availability

All data can be obtained through individual studies or by request to the authors.

## Author Contributions

conceptualization: KMM, ZR, SH, FT; methodology: KMM, SH, ATL, AB; statistical analysis: KMM, ATL, FT; writing and editing: KMM, DWB, SH, ZR, FT, REM, and others; data contribution: ZR, SH, DWB, LF, REM, MK, DLC. All authors helped with manuscript preparation and interpreted results.

## Funding: Page: 20

SH, KMM, REM, and ATL acknowledge support from 1U01AG060908. ZR, FT, and MJ acknowledge support from the National Excellence Program (126823) and the Scientific Excellence Program, TKP2020-NKA-17, at the University of Physical Education, Innovation and Technology Ministry, Hungary. REM is supported by Alzheimer’s Society major project grant AS-PG-19b-010. SRC is supported by a Sir Henry Dale Fellowship jointly funded by the Wellcome Trust, the Royal Society grant 221890/Z/20/Z.

FHS is funded by the National Institute of Health (NIH) contract N01-HC-25195 and HHSN268201500001I. The laboratory work for this investigation was funded by the Division of Intramural Research, National Heart, Lung, and Blood Institute, NIH. The analytical component of this project was funded by the Division of Intramural Research, National Heart, Lung, and Blood Institute, and the Center for Information Technology, NIH, Bethesda, MD.

JHS is supported and conducted in collaboration with Jackson State University (HHSN268201800013I), Tougaloo College (HHSN268201800014I), the Mississippi State Department of Health (HHSN268201800015I/HHSN26800001) and the University of Mississippi Medical Center (HHSN268201800010I, HHSN268201800011I and HHSN268201800012I) contracts from the National Heart, Lung, and Blood Institute and the National Institute for Minority Health and Health Disparities.

WHI program is funded by the National Heart, Lung, and Blood Institute, NIH, and the U.S. Department of Health and Human Services through contracts HHSN268201600018C, HHSN268201600001C, HHSN268201600002C, HHSN268201600003C, and HHSN268201600004C.

The LBC1921 was supported by the UK’s Biotechnology and Biological Sciences Research Council, a Royal Society–Wolfson Research Merit Award and the Chief Scientist Office (CSO) of the Scottish Government’s Health Directorates. The LBC1936 is supported by Age UK (Disconnected Mind project), the Medical Research Council (G0701120, G1001245, MR/M013111/1, MR/R024065/1), and the University of Edinburgh. Methylation typing was supported by Centre for Cognitive Ageing and Cognitive Epidemiology (Pilot Fund award), Age UK, The Wellcome Trust Institutional Strategic Support Fund, The University of Edinburgh, and The University of Queensland.

CALERIE Analysis of DNA methylation data was conducted in collaboration with Columbia University Mailman School of Public Health with support from National Institute on Aging grant R01AG061378 and non-genomic data curated as part of National Institute on Aging grant R33AG070455.

The authors wish to thank the staff, investigators, and study participants from every study for making this research possible. We would also like to thank Robert (Bobby) Brooke for reviewing and making suggestions to this manuscript.

