## Supplemental Note 1 for "DNAmFitAge: Biological Age Indicator Incorporating Physical Fitness"

*Supplemental Note 1: Other Variables*

Not all validation datasets have measurements of VO2max, FEV1, handgrip strength, or gait speed. In this case, we correlate similar fitness parameters; VO2max as a substitute for FEV1 and composite leg strength or composite physical functioning score as substitutes for gait speed. Composite leg strength is a measure of absolute peak leg flexion and extension torque, measured in Newton-meters. Composite physical functioning score combines walking and chair activities and ranges from 0 to 12 with 12 being best physical functioning. We expect VO2max, composite leg strength, and composite physical functioning to have positive correlation with their respective DNAm fitness parameter biomarker. VO2max and composite leg strength are used in CALERIE, and composite physical functioning score is used in InChianti and WHI.

LBC21 measures self-reported days per month spent exercising; participants with at most 12 days of reported exercise per month were included. LBC36 measures level of physical activity using an electronic activity monitor and then categorizes people into one of six categories: sedentary, light, low-light activity, high-light, moderate to vigorous, and vigorous activity. LBC36 participants with sedentary to low-light activity were included for analysis. JHS categorizes participants into poor, intermediate, or ideal physical activity health; participants with poor or intermediate categorization were included. InChianti measures physical functioning as a composite score from 0 to 12 with 12 being a perfect score; participants with scores at or below 11 were included. WHI measures physical functioning as a composite score from 0 to 100 with 100 being a perfect score. WHI participants with scores at or below 85 were included; outliers were also excluded using scores beyond 1.5 times the interquartile range.

DNAmPhenoAge is an estimate of epigenetic age constructed using DNAm composite clinical measures of phenotypic age [27]. DNAmPAI-1 is a surrogate marker of plasma protein plasminogen activator inhibitor level 1, and DNAmGDF-15 is a surrogate marker for growth differentiation factor 15 [28].
