## Supplemental Note 2 for "DNAmFitAge: Biological Age Indicator Incorporating Physical Fitness"

*Supplemental Note 2: Validation Datasets*

Our validation datasets involved six cohorts: the Lothian Birth Cohorts (1921 and 1936), Comprehensive Assessment of Long-term Effects of Reducing Intake of Energy (CALERIE), the Women's Health Initiative (WHI), Jackson Heart Study (JHS), and Invecchiare in Chianti, aging in the Chianti area (InChianti). Below we describe each study cohort/datasets in more detail.

### *Lothian Birth Cohorts*

The Lothian Birth Cohort (LBC) consists of two longitudinal studies evaluating cognition and brain aging of older adults who were born in either 1921 (LBC21) or 1936 (LBC36) and lived in Edinburgh or the surrounding Lothian regions. LBC21 was started in 1999 and LBC36 began in 2004. The Lothian Birth Cohort 1936 (LBC1936) was established to study cognitive aging in surviving members of the 1947 Scottish Mental Survey. Ethical approval was obtained from the Multi-Centre Ethics Committee for Scotland and Lothian Research Ethics Committee. National Records of Scotland provided regular updates on mortality data for the LBC participants via data linkage with the National Health Service Central Register.

### *LBC1921*

Participants were born in 1921 and most completed a cognitive ability test around age of 11 years in the Scottish Mental Survey 1932 (SMS1932). The SMS1932 was administered nationwide to almost all 1921-born children who attended school in Scotland in June 1932. The cognitive test was the Moray House Test No. 12. The LBC1921 study attempted to follow up individuals who might have completed the SMS1932 and resided in the Lothian region (Edinburgh and its surrounding areas) of Scotland; 550 people (N = 234, 43% men) were successfully traced and participated in the study from the age of 79 years. To date, there have been four additional follow-up waves at average ages of 83, 87, 90, and 92 years. The cohort has been studied during the later-life waves, including blood biomarkers, cognitive testing, and psycho-social, lifestyle, and health measures.

### *LBC1936*

The methylation mortality survival analysis was investigated in Lothian Birth Cohort 1936 (LBC1936). All participants were born in 1936 and most had taken part in the Scottish Mental Survey 1947. These participants attended Scottish schools in June 1947. The cognitive test administered was the same Moray House Test No. 12. A total of 1,091 participants (n = 548, 50% men) who were living in the Lothian area of Scotland were re-contacted in later life. Data has since been collected in waves at three time points.

### *LBC DNAm methylation quantification*

Whole blood DNA methylation was measured using the Illumina HumanMethylation450BeadChips from 514 whole blood samples in LBC1921 and from 1,004 samples in LBC1936. Samples were extracted at MRC Technology, Western General Hospital, Edinburgh (LBC1921) and the Wellcome Trust Clinical Research Facility (WTCRF), Western General Hospital, Edinburgh (LBC1936), using standard methods. Methylation typing of 485,512 probes was performed at the WTCRF. Raw intensity data were background-corrected and methylation beta-values generated using the R minfi package. Quality control analysis was performed to remove probes with a low (<95%) detection rate at $P$ <0.01. Manual inspection of the array control probe signals was used to identify and remove low quality samples (for example, samples with inadequate hybridization, bisulfite conversion, nucleotide extension, or staining signal). The Illumina-recommended threshold was used to eliminate samples with a low call rate (samples with <450,000 probes detected at $P$ <0.01). Since the LBC samples had previously been genotyped using the Illumina 610-Quadv1 genotyping platform, genotypes derived from the 65 SNP control probes on the methylation array using the wateRmelon package were compared to those obtained from the genotyping array to ensure sample integrity. Samples with a low match of genotypes with SNP control probes, which could indicate sample contamination or mix-up, were excluded (n = 9). Moreover, eight subjects whose predicted sex, based on XY probes, did not match reported sex were also excluded.

***CALERIE***

Comprehensive Assessment of Long-term Effects of Reducing Intake of Energy (CALERIE) was a Phase 2 clinical trial started in 2007 studying young to middle-aged healthy adults [13]. CALERIE is the first clinical trial to focus on the effects of sustained CR in humans. It was completed in May 2013 as a two-year three-site randomized controlled trial in young and middle-aged non-obese healthy men and women (N = 220). Participants were randomized in a 2:1 fashion to 25% caloric restriction (CR) or ad libitum control group (diet is available at all times). All participants needed to have a baseline body mass index (BMI) of 22-27.9 kg/m$^2$ (lean to slightly overweight). Each participant has 1) behavioral counselor (Masters of doctoral in psychology) AND 2) registered dietician who follow with them for the whole 2 years. 25% reduction and caloric goals are calculated based on each person's initial food intake at baseline. They must meet with the dietician 2-3 times a week and record food intake. Two consecutive 14-day doubly labeled water studies are conducted with each participant at baseline with the average used to determine AL TEE (total energy expenditure); from this, the 25% CR prescription for that participant is derived. An average of 12% caloric reduction was achieved in the CR group throughout the study.

***CALERIE DNAm methylation quantification***

DNA methylation was measured from Illumina EPIC 850k Arrays (Illumina Inc., San Diego, CA) as per the manufacturer's protocol. DNA methylation was derived from whole blood samples. CALERIE methylation assays were run by the Molecular Genomics Shared Resource at Duke Molecular Physiology Institute, Duke University (USA). Quality control of sample handling included comparison of clinically reported sex versus sex of the same samples determined by analysis of methylation levels of CpG sites on the X chromosome. Methylation beta values were generated using the Bioconductor *minfi* package with Noob background correction.

CALERIE data are available at https://calerie.duke.edu/samples-data-access-and-analysis.

***Women's Health Initiative***

The WHI is a national study that enrolled postmenopausal women aged 50-79 years into the clinical trials (CT) or observational study (OS) cohorts between 1993 and 1998[4, 5]. We included 4,079 WHI participants with available phenotype and DNA methylation array data:

2,107 women from “*Broad Agency Award 23*” (WHI BA23). WHI BA23 focuses on identifying miRNA and genomic biomarkers of coronary heart disease (CHD), integrating the biomarkers into diagnostic and prognostic predictors of CHD and other related phenotypes, and other objectives can be found in https://www.whi.org/researchers/data/WHIStudies/StudySites/BA23/Pages/home.aspx

The total number of age-related conditions was based on Alzheimer’s disease, amyotrophic lateral sclerosis, arthritis, cancer, cataract, CVD, glaucoma, emphysema, hypertension, and osteoporosis.

***WHI DNA methylation quantification***

Bisulfite conversion using the Zymo EZ DNA Methylation Kit (Zymo Research, Orange, CA, USA) as well as subsequent hybridization of the HumanMethylation450k Bead Chip (Illumina, San Diego, CA), and scanning (iScan, Illumina) were performed according to the manufacturers protocols by applying standard settings. DNA methylation levels (β values) were determined by calculating the ratio of intensities between methylated (signal A) and un-methylated (signal B) sites. Specifically, the β value was calculated from the intensity of the methylated (M corresponding to signal A) and un-methylated (U corresponding to signal B) sites, as the ratio of fluorescent signals $\beta = Max(M,0)/[Max(M,0)+Max(U,0)+100]$. Thus, β values range from 0 (completely un-methylated) to 1 (completely methylated).

***Jackson Heart Study***

The JHS is a large, population-based observational study evaluating the etiology of cardiovascular, renal, and respiratory diseases among African Americans residing in the three counties (Hinds, Madison, and Rankin) that make up the Jackson, Mississippi metropolitan area. The age at enrollment for the unrelated cohort was 35-84 years; the family cohort included related individuals >21 years old. Participants provided extensive medical and social history, had an array of physical and biochemical measurements and diagnostic procedures, and provided genomic DNA during a baseline examination (2000-2004) and two follow-up examinations (2005-2008 and 2009-2012). Annual follow-up interviews and cohort surveillance are ongoing. In our analysis, we used the visits at baseline from 1747 individuals as part of project JHS ancillary study ASN0104, available with both phenotype and DNA methylation array data. Total numbers of age-related conditions were based on hypertension, type 2 diabetes, kidney dysfunction based on ever dialysis, and CVD.

***JHS DNA methylation quantification***

Peripheral blood samples were collected at the baseline. DNA was extracted using the Gentra Puregene blood kit (Gentra System, MN, Minnesota, USA). Methylation beta values were generated using the Bioconductor *minfi* package with Noob background correction.

***Invecchiare in Chianti, aging in the Chianti area (InChianti)***

The InChianti (Invecchiare in Chianti, aging in the Chianti area) cohort is a representative population-based study of older persons enrolling individuals aged 20 years and older from two areas in the Chianti region of Tuscany, Italy, http://inchiantistudy.net/wp/. One major goal of the study is to translate epidemiological research into geriatric clinical tools, ultimately advancing clinical applications in older persons. Of the cohort, 924 observations from 484 individuals with both phenotype information and DNA methylation data were including in our studies. The observations were collected from baseline in 1998 and the third follow-up visit in 2007. All participants provided written informed consent to participate in this study. The study complied with the Declaration of Helsinki. The Italian National Institute of Research and Care on Aging Institutional Review Board approved the study protocol. We computed the total number of age-related conditions based on cancer, hypertension, myocardial infarction, Parkinson's disease, stroke and type 2 diabetes.

***InChianti DNA methylation quantification***

Genomic DNA was extracted from buffy coat samples using an AutoGen Flex and quantified on a Nanodrop1000 spectrophotometer prior to bisulfite conversion. Blood DNA methylation was taken twice over the span of nine years in a total of 966 people. Genomic DNA was bisulfite converted using Zymo EZ-96 DNA Methylation Kit (Zymo Research Corp., Irvine, CA) as per the manufacturer's protocol. CpG methylation status of 485,577 CpG sites was determined using the Illumina Infinium HumanMethylation450 BeadChip (Illumina Inc., San Diego, CA) as per the manufacturer's protocol and as previously described [11]. Initial data analysis was performed using GenomeStudio 2011.1 (Model M Version 1.9.0, Illumina Inc.). Threshold call rate for inclusion of samples was 95%. Quality control of sample handling included comparison of clinically reported sex versus sex of the same samples determined by analysis of methylation levels of CpG sites on the X chromosome. Methylation beta values were generated using the Bioconductor *minfi* package with Noob background correction.
