## Supplementary figures and images for "DNAmFitAge: Biological Age Indicator Incorporating Physical Fitness"

### Supplemental Table 1

#### Slide 1
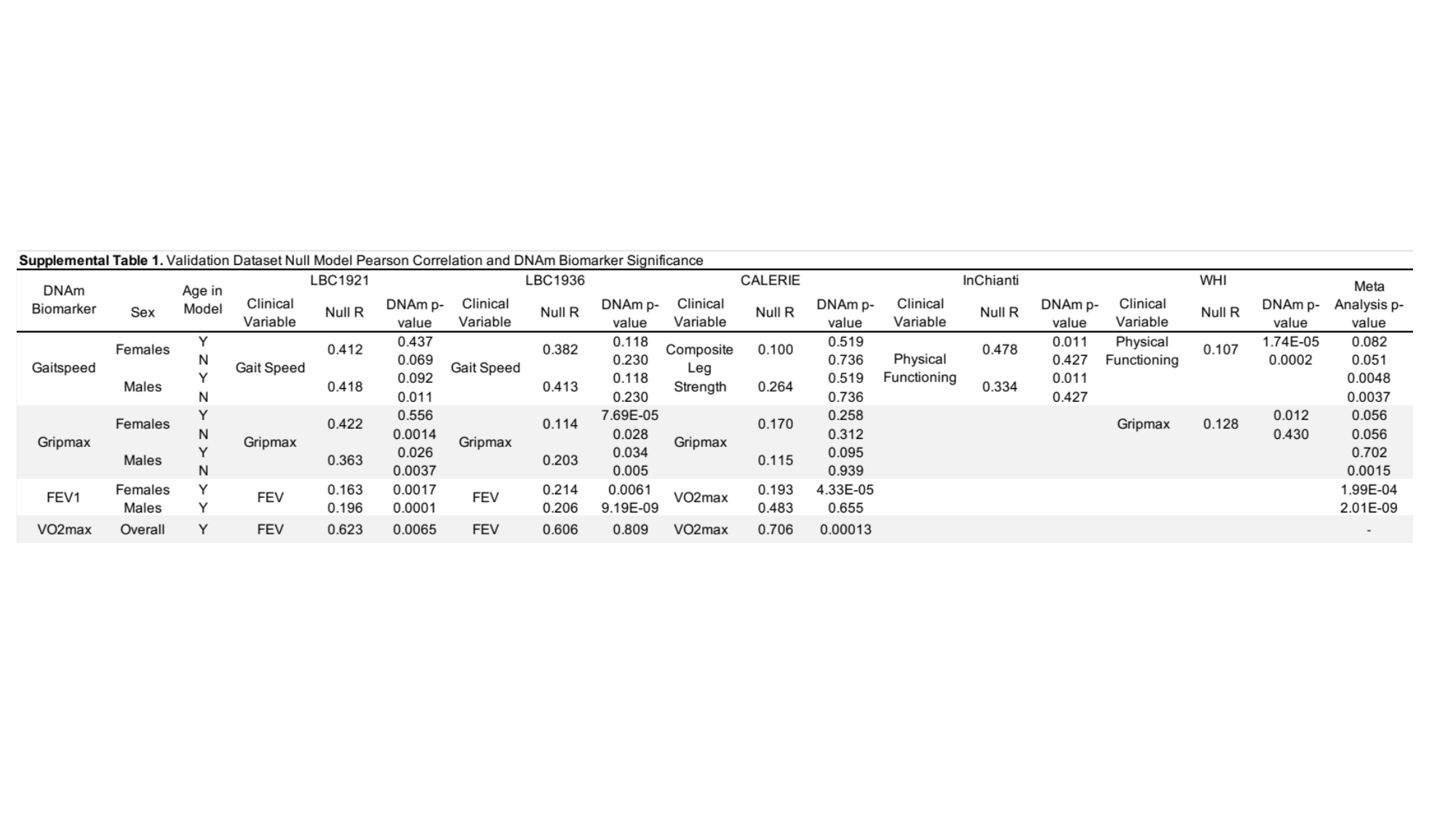

### Supplemental Table 2

#### Slide 1
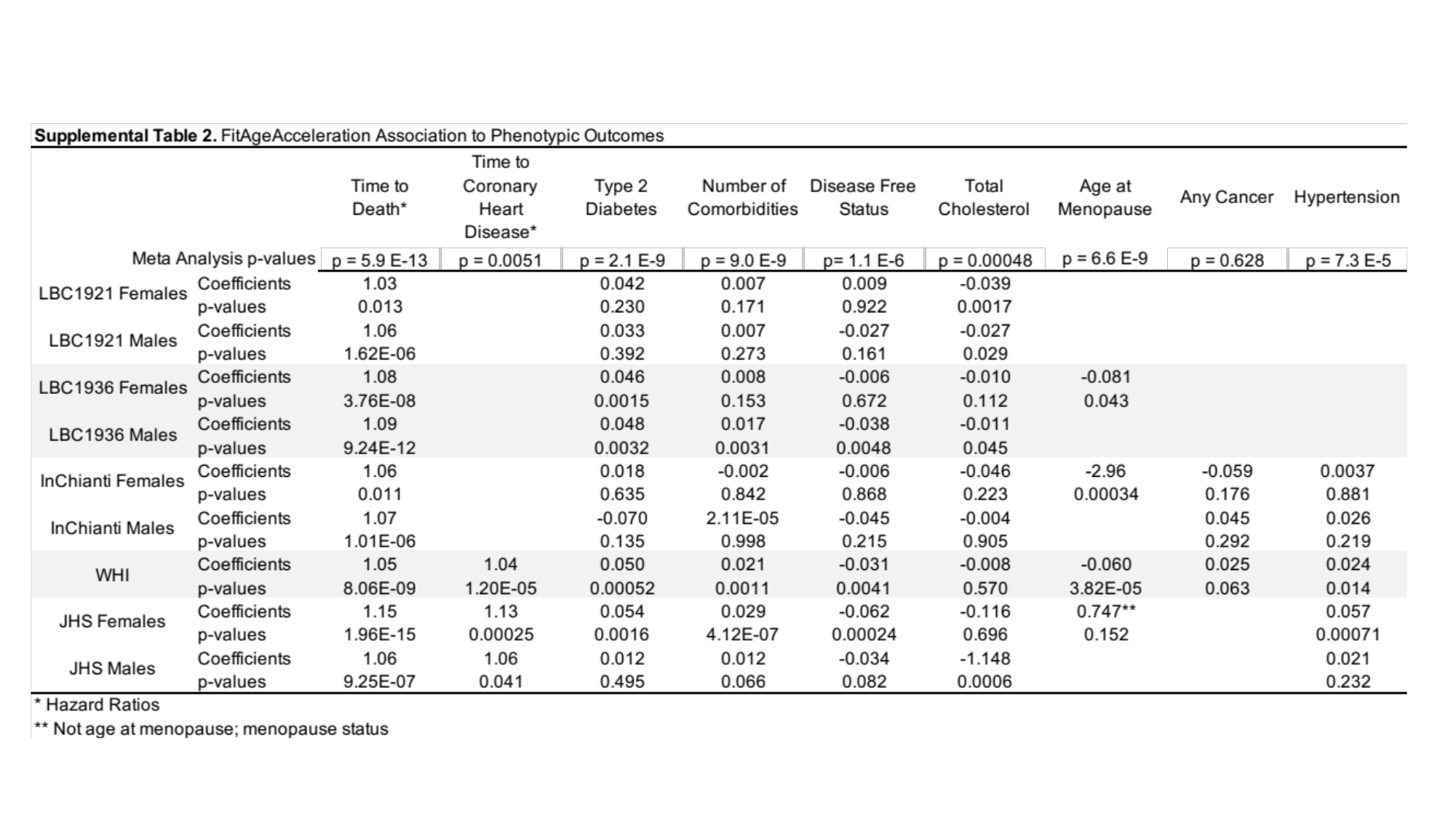

### Supplemental Table 3

#### Slide 1
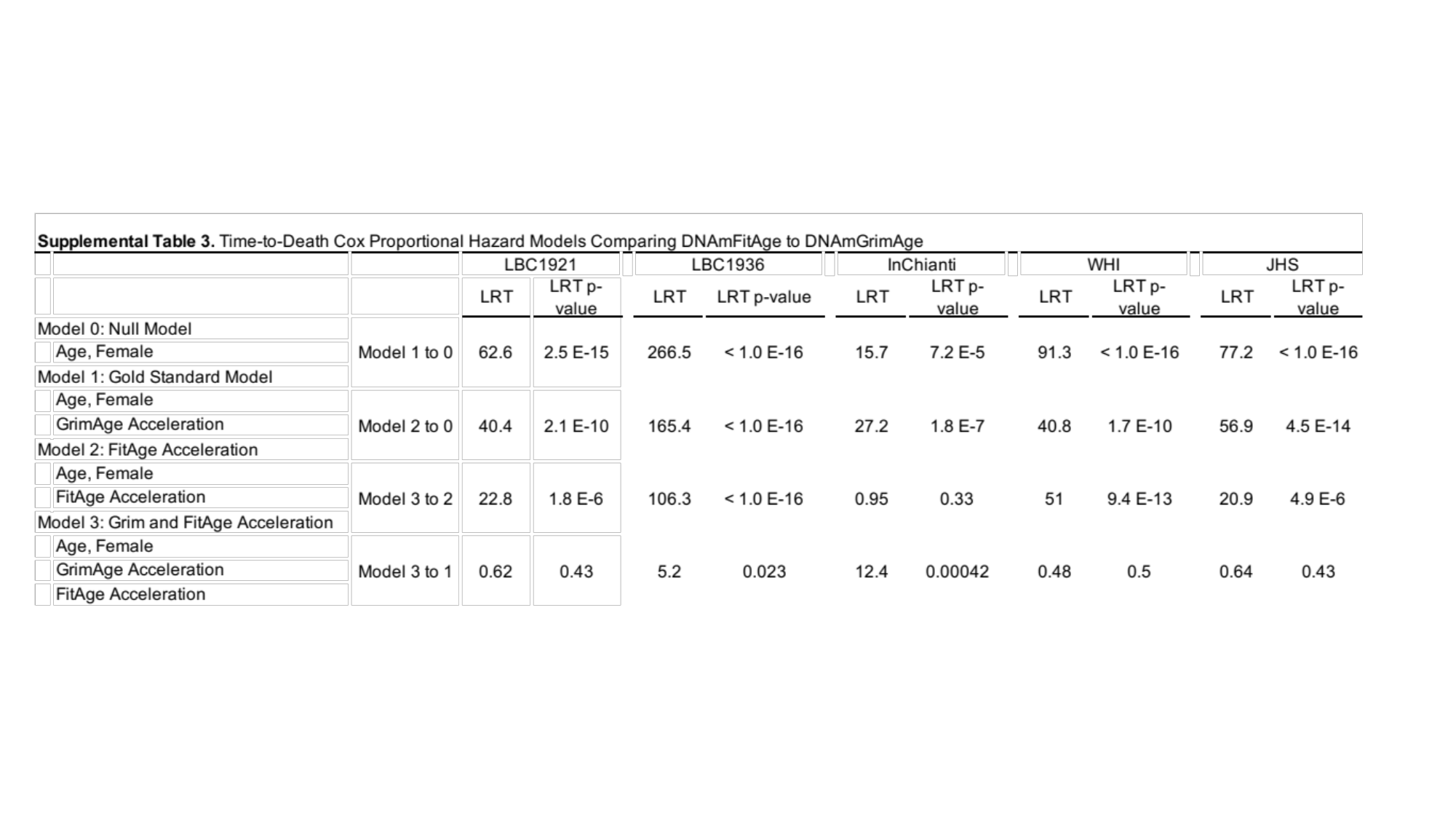
