## Supplemental Figure 1 for "DNAmFitAge: Biological Age Indicator Incorporating Physical Fitness"

### Slide 1
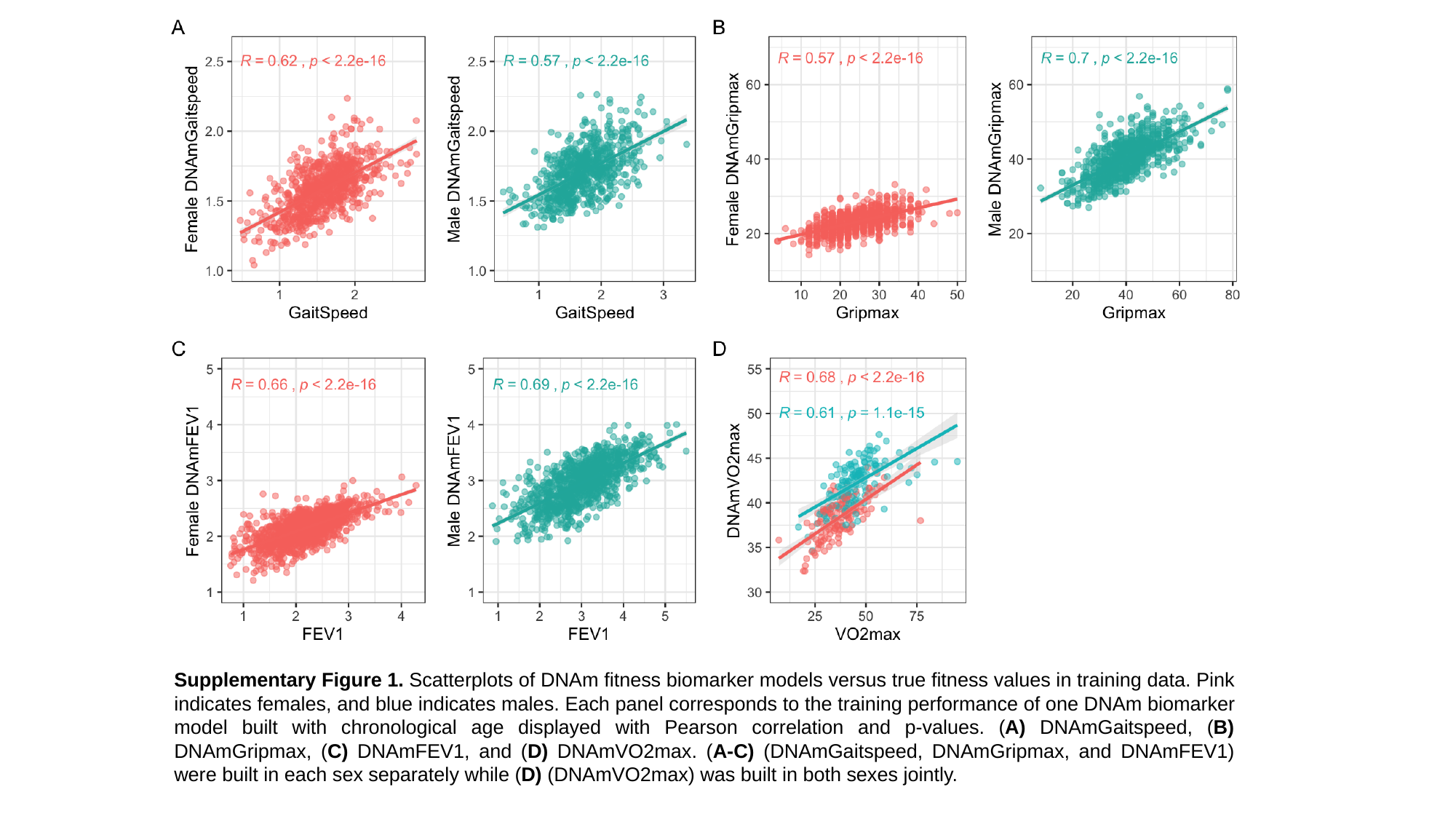

Supplementary Figure 1. Scatterplots of DNAm fitness biomarker models versus true fitness values in training data. Pink indicates females, and blue indicates males. Each panel corresponds to the training performance of one DNAm biomarker model built with chronological age displayed with Pearson correlation and p-values. (A) DNAmGaitspeed, (B) DNAmGripmax, (C) DNAmFEV1, and (D) DNAmVO2max. (A-C) (DNAmGaitspeed, DNAmGripmax, and DNAmFEV1) were built in each sex separately while (D) (DNAmVO2max) was built in both sexes jointly.
