## Supplemental Figure 2 for "DNAmFitAge: Biological Age Indicator Incorporating Physical Fitness"

### Slide 1
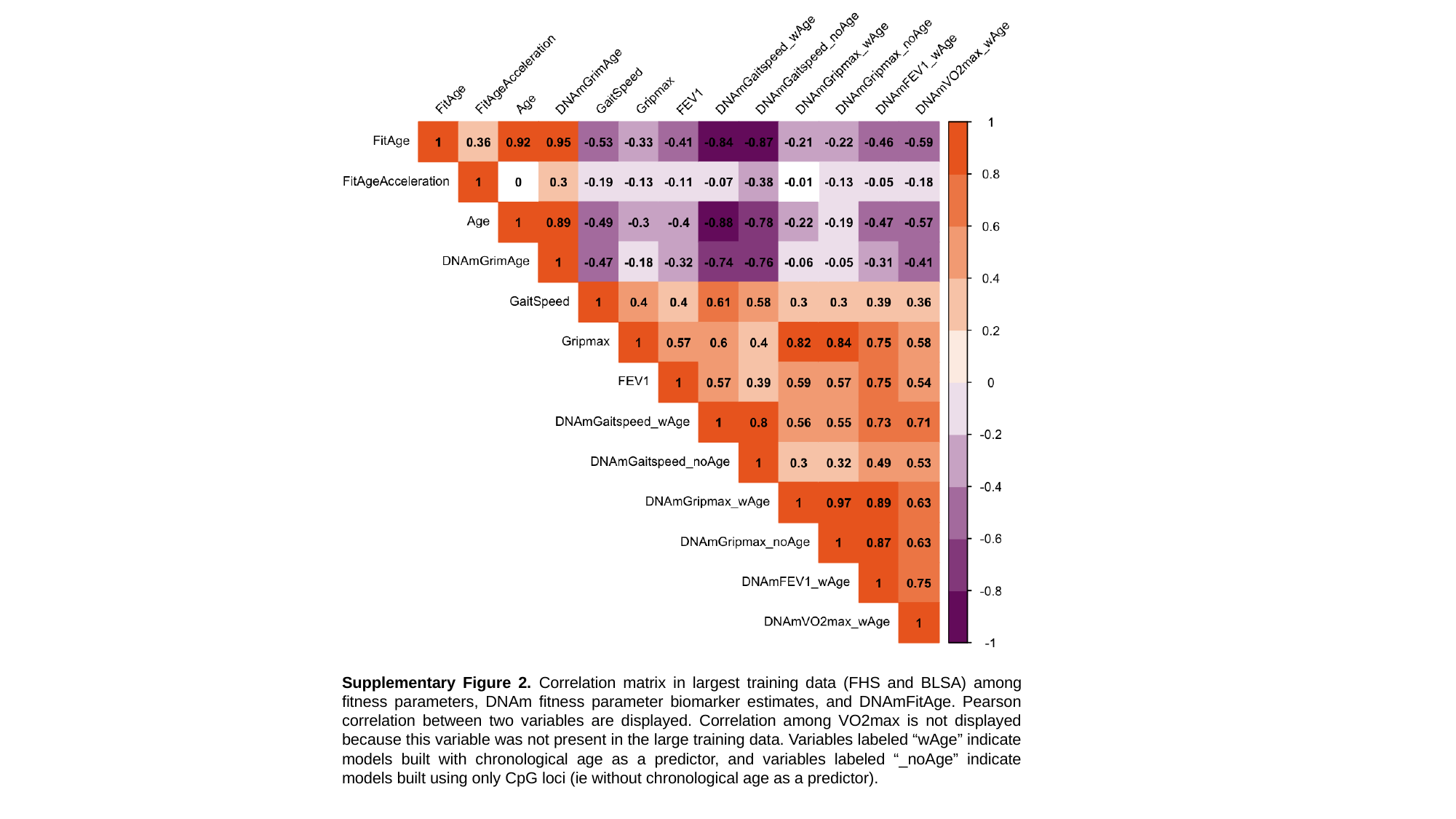

Supplementary Figure 2. Correlation matrix in largest training data (FHS and BLSA) among fitness parameters, DNAm fitness parameter biomarker estimates, and DNAmFitAge. Pearson correlation between two variables are displayed. Correlation among VO2max is not displayed because this variable was not present in the large training data. Variables labeled “wAge” indicate models built with chronological age as a predictor, and variables labeled “_noAge” indicate models built using only CpG loci (ie without chronological age as a predictor).
