## Supplemental Figure 3 for "DNAmFitAge: Biological Age Indicator Incorporating Physical Fitness"

### Slide 1
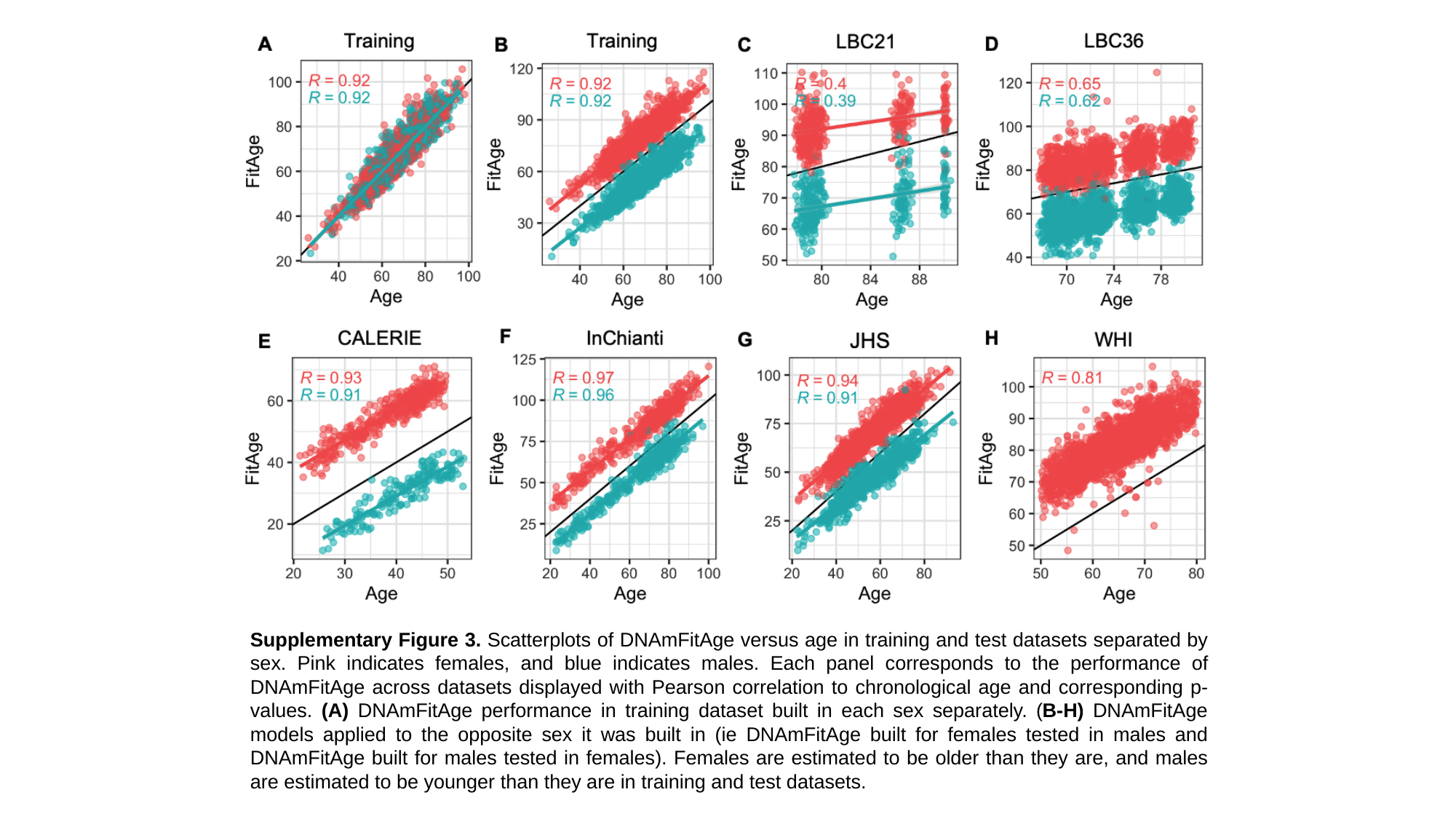

Supplementary Figure 3. Scatterplots of DNAmFitAge versus age in training and test datasets separated by sex. Pink indicates females, and blue indicates males. Each panel corresponds to the performance of DNAmFitAge across datasets displayed with Pearson correlation to chronological age and corresponding p-values. (A) DNAmFitAge performance in training dataset built in each sex separately. (B-H) DNAmFitAge models applied to the opposite sex it was built in (ie DNAmFitAge built for females tested in males and DNAmFitAge built for males tested in females). Females are estimated to be older than they are, and males are estimated to be younger than they are in training and test datasets.
