## Supplemental Figure 4 for "DNAmFitAge: Biological Age Indicator Incorporating Physical Fitness"

### Slide 1
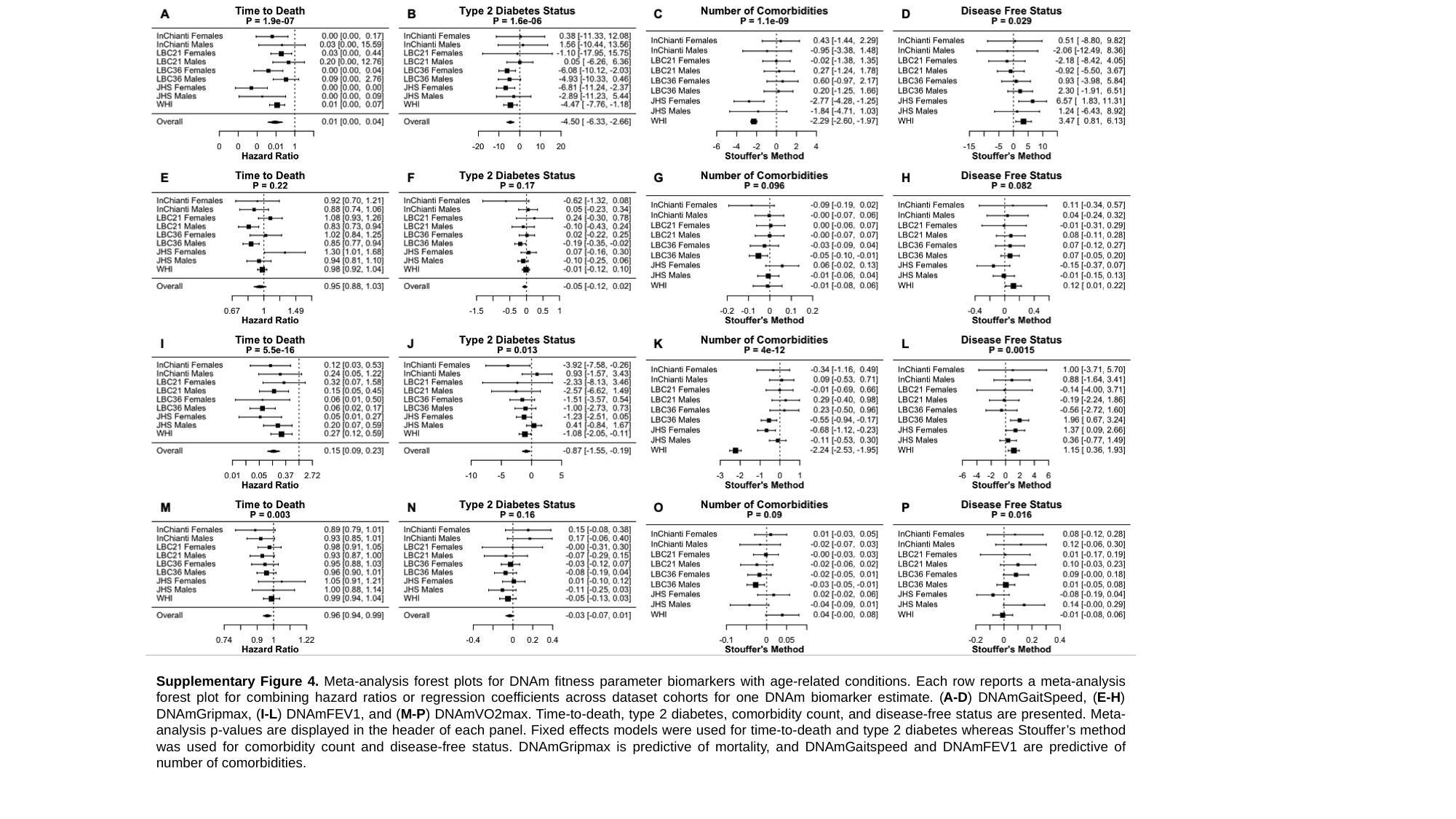

Supplementary Figure 4. Meta-analysis forest plots for DNAm fitness parameter biomarkers with age-related conditions. Each row reports a meta-analysis forest plot for combining hazard ratios or regression coefficients across dataset cohorts for one DNAm biomarker estimate. (A-D) DNAmGaitSpeed, (E-H) DNAmGripmax, (I-L) DNAmFEV1, and (M-P) DNAmVO2max. Time-to-death, type 2 diabetes, comorbidity count, and disease-free status are presented. Meta-analysis p-values are displayed in the header of each panel. Fixed effects models were used for time-to-death and type 2 diabetes whereas Stouffer’s method was used for comorbidity count and disease-free status. DNAmGripmax is predictive of mortality, and DNAmGaitspeed and DNAmFEV1 are predictive of number of comorbidities.
